# Patient-derived cells demonstrate that mutated RyR2 calcium leak underlies autism spectrum disorder and inherited arrhythmias

**DOI:** 10.1101/2025.07.26.25332119

**Authors:** Sarah Colombani, Hugo Benoit, Marco C. Miotto, Steve Reiken, Albin A. Bernardin, Florence Bernex, Romain Desprat, Marie Vincenti, Virginie Andry, Yannick Goumon, Andrew R. Marks, Jean-Luc Pasquié, Alain Lacampagne, Albano C. Meli

## Abstract

Catecholaminergic polymorphic ventricular tachycardia (CPVT) and autism spectrum disorder (ASD) are increasingly recognized as comorbid conditions, yet their shared molecular mechanisms remain unclear. This study investigates a novel RyR2-R169P mutation identified in a patient diagnosed with both CPVT and ASD, hypothesizing that this mutation drives calcium (Ca^2+^) dysregulation in cardiac and neuronal cells.

Using patient-derived induced pluripotent stem cells, we generated ventricular-like cardiomyocytes and midbrain neurons. In cardiomyocytes, the RyR2-R169P mutation increased diastolic Ca^2+^ leak, elevated single-channel open probability, and induced arrhythmogenic Ca^2+^ waves under β-adrenergic stress. Similarly, neurons exhibited abnormal cytosolic Ca^2+^ levels, enlarged soma size, and a clear trend to disrupted neurotransmitter release, including reduced GABA and elevated L-DOPA and serotonin. RyR2 biochemical analysis showed reduced phosphorylation of RyR2 by CaMKII, increased PKA dependent phosphorylation and dissociation of calstabin2 in neurons.

Pharmacological stabilization of RyR2 with S107 normalized Ca^2+^ handling in both cell types, restored neuronal morphology, and prevented calstabin2 depletion and CaMKII phosphorylation increase. S107 restores normal neurotransmitter release only when treatment starts before neuronal differentiation. Structural modeling revealed that the R169P mutation destabilizes the N-terminal domain of RyR2, priming the channel for pathological Ca^2+^ leak.

These findings establish RyR2-R169P as a dual regulator of Ca^2+^ homeostasis, directly linking cardiac arrhythmogenesis to neurodevelopmental deficits. Our results highlight RyR2 dysfunction as a shared mechanism in CPVT-ASD comorbidity and propose Rycals as a promising therapeutic candidate for mitigating Ca^2+^-driven pathologies in both tissues. This work demonstrates the importance of RyR2 functional integrity in neurodevelopmental processes.

**One Sentence Summary:** Novel RyR2-R169P mutation causes calcium leak in hiPSC-derived cardiomyocytes and neurons, linking CPVT to autism via RyR2 dysfunction.

## Introduction

Catecholaminergic polymorphic ventricular tachycardia (CPVT) is a life-threatening cardiac arrhythmia syndrome characterized by adrenergic-induced ventricular tachycardia in structurally normal hearts, often manifesting as syncope or sudden cardiac death in children and adolescents ^1^. Over 80% of CPVT cases are linked to mutations in the type 2 ryanodine receptor (RyR2), the primary calcium (Ca^2+^) release channel of the sarcoplasmic reticulum in cardiomyocytes ^2^. Dysfunctional RyR2 channels cause diastolic Ca^2+^ leak, triggering delayed afterdepolarizations and arrhythmias via sodium-calcium exchanger activation ^3^. While murine models and patient-derived induced pluripotent stem cell cardiomyocytes (hiPSC-CMs) have elucidated cardiac mechanisms ^4, 5^, emerging clinical observations reveal that CPVT patients frequently exhibit comorbid neuropsychiatric disorders, including autism spectrum disorder (ASD) ^6^, suggesting a broader role for RyR2 dysfunction beyond the heart.

ASD, a neurodevelopmental condition marked by impaired social communication and repetitive behaviors, has been increasingly associated with dysregulated intracellular Ca^2+^ signaling ^7^. RyR2 is also expressed in neurons, where it modulates Ca^2+^ dynamics critical for synaptic plasticity, neurotransmitter release, and neural network formation ^8^. Notably, murine models harboring CPVT-associated RyR2 mutations exhibit not only cardiac arrhythmias but also neurobehavioral deficits, including seizures and anxiety-like behaviors ^9^. Human studies further link RyR2 dysregulation to neurodegenerative and neuropsychiatric conditions, implicating Ca^2+^ mishandling in diverse central nervous system (CNS) pathologies ^10^. However, whether specific RyR2 mutations can directly drive both cardiac and neurodevelopmental phenotypes in humans remains unresolved.

Here, we report a novel RyR2-R169P mutation identified in a patient diagnosed with CPVT and ASD. This case provides a unique opportunity to investigate how a single genetic lesion disrupts Ca^2+^ homeostasis across tissues. We hypothesized that RyR2-R169P induces pathological Ca^2+^ leak in both cardiomyocytes and neurons, coupling arrhythmogenesis to neurodevelopmental dysfunction. To test this, we generated patient-derived hiPSCs, differentiated them into ventricular-like cardiomyocytes (hiPSC-CMs) and CNS neurons (hiPSC-NRs), and compared their Ca^2+^ handling properties to controls. Structural modeling and pharmacological interventions further dissected the mutation’s mechanistic impact.

Our findings reveal that RyR2-R169P destabilizes the channel’s N-terminal domain, predisposing it to Ca^2+^ leak under stress. In hiPSC-CMs, this leak drives arrhythmogenic Ca^2+^ waves, while in hiPSC-NRs, it disrupts neuronal Ca^2+^ transients, soma morphology, and neurotransmitter balance. Crucially, the RyR2 stabilizer S107 rescues these defects, highlighting a shared therapeutic target for cardiac and neuronal pathologies. This study establishes RyR2-R169P as a dual regulator of Ca^2+^ homeostasis, directly linking CPVT to ASD through cell-autonomous mechanisms, and underscores the value of hiPSC models in dissecting complex genotype-phenotype relationships.

## Material and methods

### Ethics statement

Written informed consent form was obtained from the parents of the minor patient with the RyR2-R169P mutation, who agreed to provide blood samples for hiPSC generation and basic research. This study was conducted in accordance with the Declaration of Helsinki and was approved by the Montpellier Hospital Review Board Committee (approval number 1003-HPS2). The male healthy control and RyR2-H29D and its isogenic control hiPSC lines used in this study were previously characterized ^5, 11^.

### 3D in silico modeling

3D *in silico* modeling was performed using the structures of human RyR2 in the closed (PDB: 7UA5) and open (PDB: 7UA9) states and of the mouse RyR2-R176Q (PDB: 6WOU). Visualization, *in silico* mutation, and hydrogen bond analysis was performed using the software ChimeraX ^12^.

### hiPSC generation and maintenance

Following blood sample and PBMC isolation, we reprogrammed in bulk and fully characterized the hiPSC line from the CPVT patient harboring RyR2-R169P as previously described ^13^. The hiPSC were maintained on extracellular matrix Matrigel hES-Qualified and cultured into StemFlex culture media at 37°C in 5% O_2_ and 5% CO_2_.

### Cardiac differentiation protocol

We carried out cardiac differentiation using the 2D cardiac sheet protocol, which modulates Wnt/β-catenin signaling pathway, using CHIR99021 and Wnt-C59 cytokines, as we have previously published ^5^. At day 20, the media RPMI/B27 was supplemented with 100nM of Triiodothyronine (T3) and 1µM of Dexamethasone to improve the ventricular-like hiPSC-CM phenotype ^14^. All experiments were done at day 30 of the cardiac differentiation.

A detailed Methods section can be found in the Supplementary material section.

## Results

### Clinical characterization identified ASD and CPVT

We present the case of a male child born in the late 2000, followed for neurodevelopmental disorders. He was diagnosed with dysphasia and coordination disorder (dyspraxia). He was born at term via cesarean section with normal Apgar scores and birth measurements. He experienced delayed motor milestones, including late walking and difficulty with running and coordination in early childhood. He exhibited behaviors that regularly put him in danger and did not form sentences, only making word associations. Speech therapy began in 2013. He received educational support and was later enrolled in a special needs program.

A psychometric assessment conducted during early schooling (WISC-5 score) ^15^, which found a Verbal Comprehension Index (VCI) of 72, a Perceptual Reasoning Index (PRI) of 69, a Working Memory Index (WMI) of 50, and a Processing Speed Index (PSI) of 66, suggesting a mild intellectual disability. Attention difficulties were also noted in the psychometric assessment. The patient did not experience epileptic seizures at any point during clinical evaluation, and electroencephalography recordings revealed no epileptiform activity or other electrophysiological abnormalities in the CNS. These findings exclude seizure-related electrophysiological dysfunction as a component of the patient’s neurological phenotype. The neurological studies carried out on this patient led to the diagnosis of an autism spectrum disorder (ASD).

In 2016, he experienced a pool incident with loss of consciousness while playing with a family member. He was immediately pulled from the water, with a noted cardiopulmonary arrest and resuscitation initiated by a volunteer firefighter present, leading to a gradual return of consciousness. Defibrillation was not needed. Upon EMS physician arrival, the ECG showed bidirectional ventricular tachycardia (VT), which spontaneously resolved (Fig. 1A), allowing for the clinical diagnosis of CPVT. Upon admission to Intensive Care, his weight and height were within the expected range for his age group. The ECG then showed sinus bradycardia at 47/min with narrow QRS complexes and a QTc of 420ms (Fig. 1B). Cardiac ultrasound noted marked apical trabeculations of the left ventricle without segmental or global kinetic disturbances. Cardiac MRI found similar anomalies without late gadolinium enhancement. Neurologically, upon admission, the brain CT and EEG were normal. Brain MRI showed no FLAIR signal abnormalities in the infra- or supratentorial regions. There was no vascular leukopathy or evidence of ischemic sequelae. Diffusion sequences showed no lesions indicative of recent ischemia.

**Figure 1:**
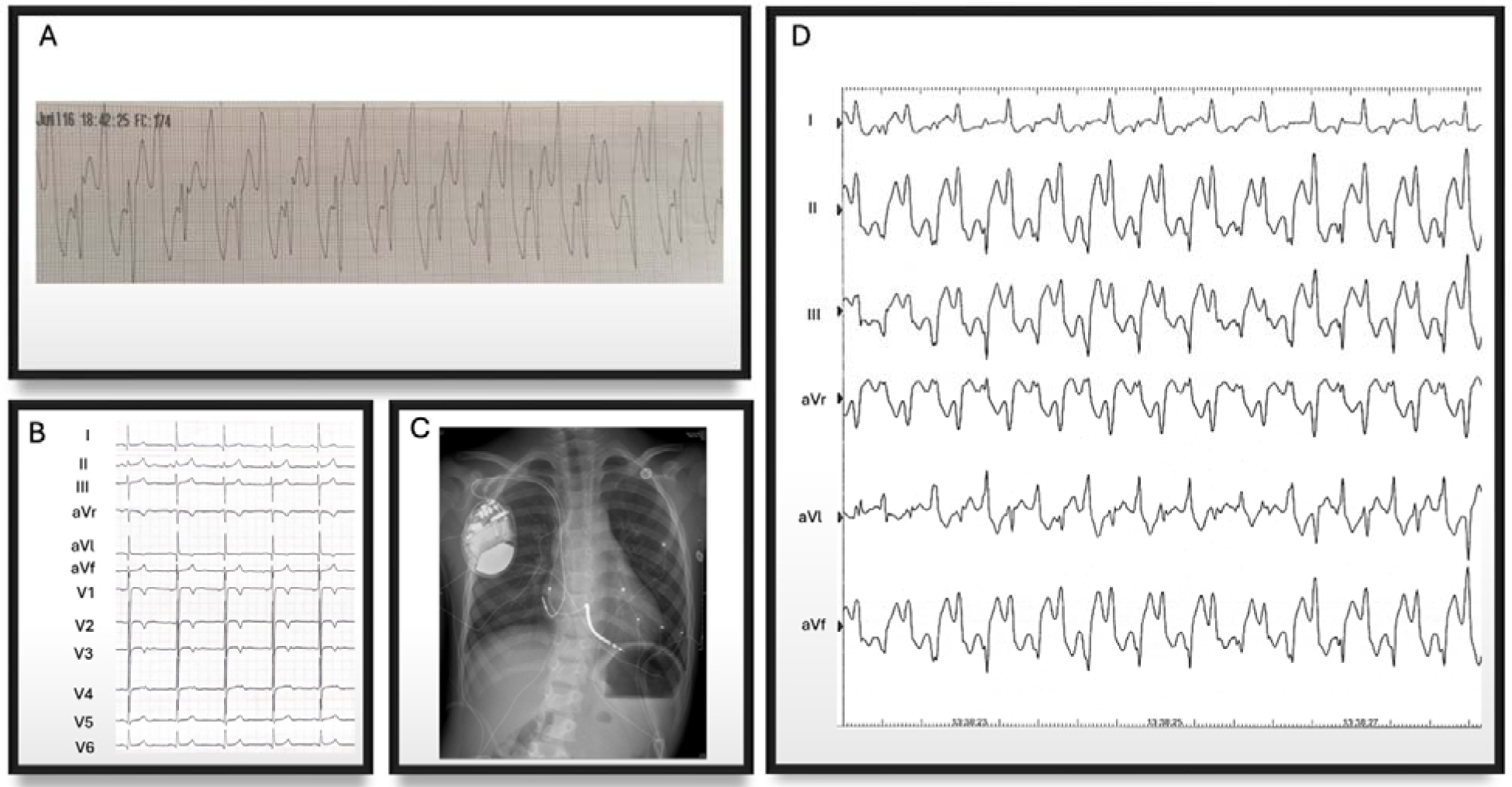
Clinical presentation. **(A)** One-lead ECG tracing of the child immediately after a near-drowning incident. This tracing shows wide-QRS tachycardia with opposite QRS axis ½ beats i.e. evidence of bidirectional tachycardia. **(B)** Baseline ECG. Sinus rhythm with normal PR interval, narrow QRS complexes, negative T waves from V1 to V3. **(C)** Chest X-Ray after dual chamber ICD implantation. **(D)** Bidirectional ventricular tachycardia induced by isoproterenol IV infusion.

We conducted pharmacological tests a week later. Under isoproterenol, atrial fibrillation (AF) appeared, followed by rapid polymorphic VT, which then organized into stable bidirectional VT (Fig. 1D and S1AB). The iv-injection of 0.5 mg of propranolol instantly stopped the VT, unmasking slow AF, which ceased after 10 seconds with a return to sinus rhythm (Fig. S1CD). A dual-chamber implantable cardioverter-defibrillator (ICD) was placed via the endovascular route (Fig. 1C), and treatment with nadolol was initiated at an initial dose of 75mg/m^2^/day. To further confirm the diagnosis of CPVT, a genetic analysis was carried out and revealed that the amino acid arginine (R) in position 169 is replaced by a proline (P) (i.e. RyR2-R169P). The FMR1 gene analysis was normal. The RyR2 mutation is also present in his asymptomatic mother in a mosaic form.

### RyR2-R169P mutation induces abnormal cytosolic Ca^2+^ handling in midbrain hiPSC-NRs

Inositol 1,4,5-trisphosphate receptors (IP_3_R) and RyR contribute to the intricate regulation of intracellular Ca^2+^ handling, which is vital for neuronal signaling and function. They influence neuronal excitability and plasticity. Recent research has shown a significant link between ASD and an abnormal Ca^2+^ handling in neurons ^7^. We investigated whether the RyR2-R169P mutation impacts the CNS by differentiating hiPSCs into midbrain hiPSC-NRs. We initially monitored intracellular Ca^2+^ handling using non-ratiometric fluorescent dyes (Fluo4-AM) to quickly sense rapid Ca^2+^ dynamics in control conditions. Healthy control (HC) hiPSC-NRs exhibited spontaneous Ca^2+^ releases of variable intensity, whereas RyR2-R169P hiPSC-NRs showed Ca^2+^ releases of higher intensity and a faster rising phase (Fig. 2A, B and D). To determine if these differences were due to leaky RyR2-R169P, we tested S107, an experimental compound known to specifically bind RyR and stabilize its closed conformation ^16^. S107 treatment further increased the mean Ca^2+^ release amplitude and did not prevent the higher Ca^2+^ release velocity (Fig. 2A, B and D). No differences were observed in decay time and frequency of the Ca^2+^ release events (Fig. 2A, C and E).

**Figure 2:**
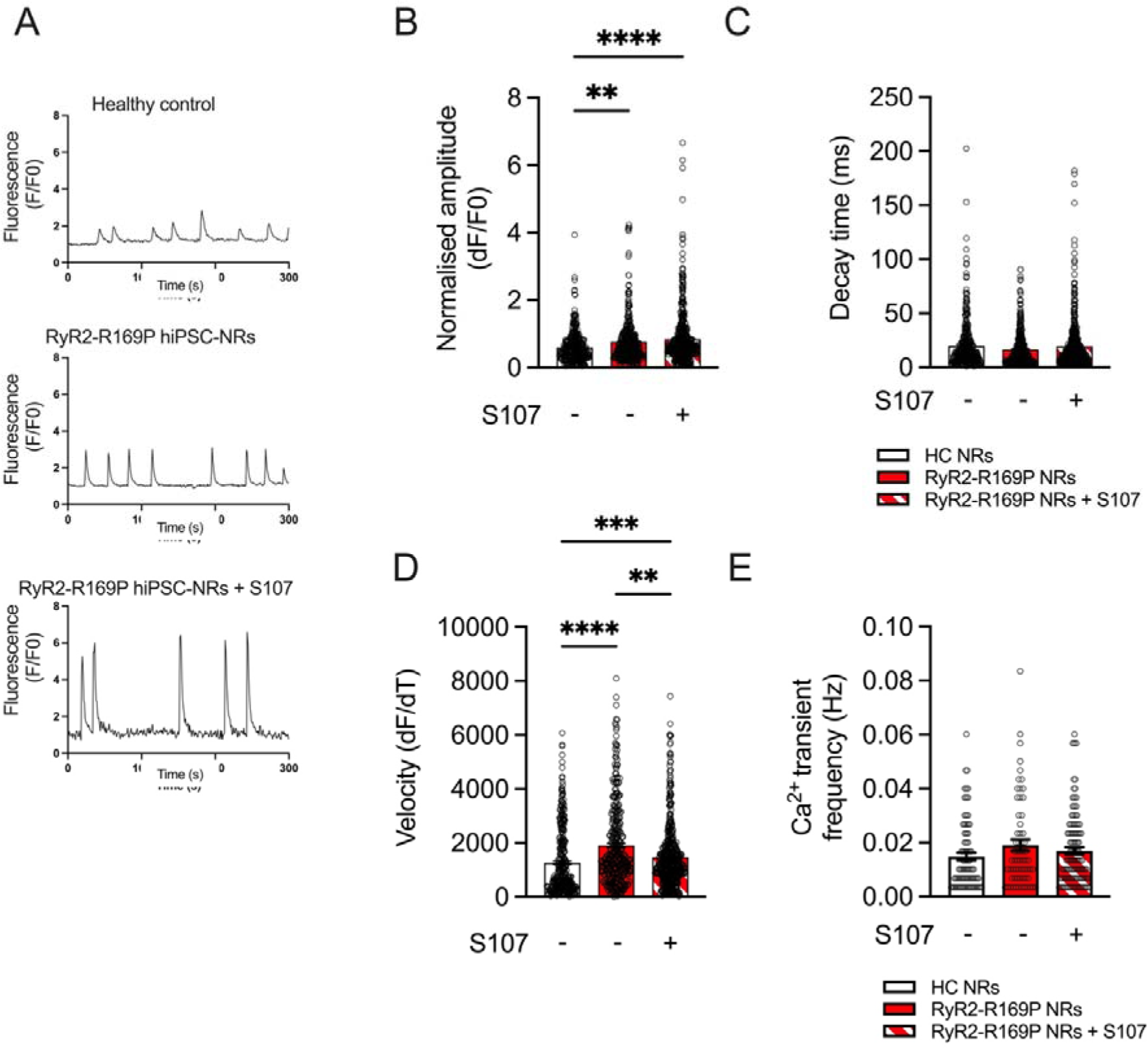
Characterization of the intracellular Ca^2+^ cycling in HC and CPVT hiPSC-NRs. **(A)** Original tracing of Ca^2+^ transients in HC and RyR2-R169P hiPSC-NRs +/- S107 (5μM). **(B)** Scatter plot showing the maximal Ca^2+^ transient amplitude in HC and RyR2-R169P hiPSC-NRs +/- S107. **(C)** Ca^2+^ reuptake time (decay time in ms); **(D)** Velocity (dF/dT) translating the maximal slope of increasing fluorescence intensity with time. **(E)** Ca^2+^ transient frequency (Hz).

### RyR2-R169P mutation induces morphological abnormalities and cytosolic Ca^2+^ leak in midbrain hiPSC-NRs

ASD has been associated with abnormal neuronal growth and migration. This includes the presence of heterotopias, which are clusters of neurons located in unusual areas of the brain ^17^. We tested whether the hiPSC-NRs harboring RyR2-R169P mutation exhibit morphological defects. Immunofluorescent experiments showed that both HC and RyR2-R169P hiPSC-NRs expressed β3-tubulin (Fig. 3A and B). We evaluated the soma and nuclear areas and found that RyR2-R169P hiPSC-NRs exhibited a larger soma (17691±2791 pixels for HC vs 27130±4576 pixels for RyR2-R169P, *p*<0.01) but a regular nucleus (Fig. 3A, B, C). Treatment with S107 (5µM overnight) reduced the soma size to HC values (Fig. 3C). A comparable number of neuronal extensions were observed both in the progenitor state (neuronal progenitors or hiPSC-NPs, Fig. S2A) and in the mature state (hiPSC-NRs) (Fig. S2B).

**Figure 3:**
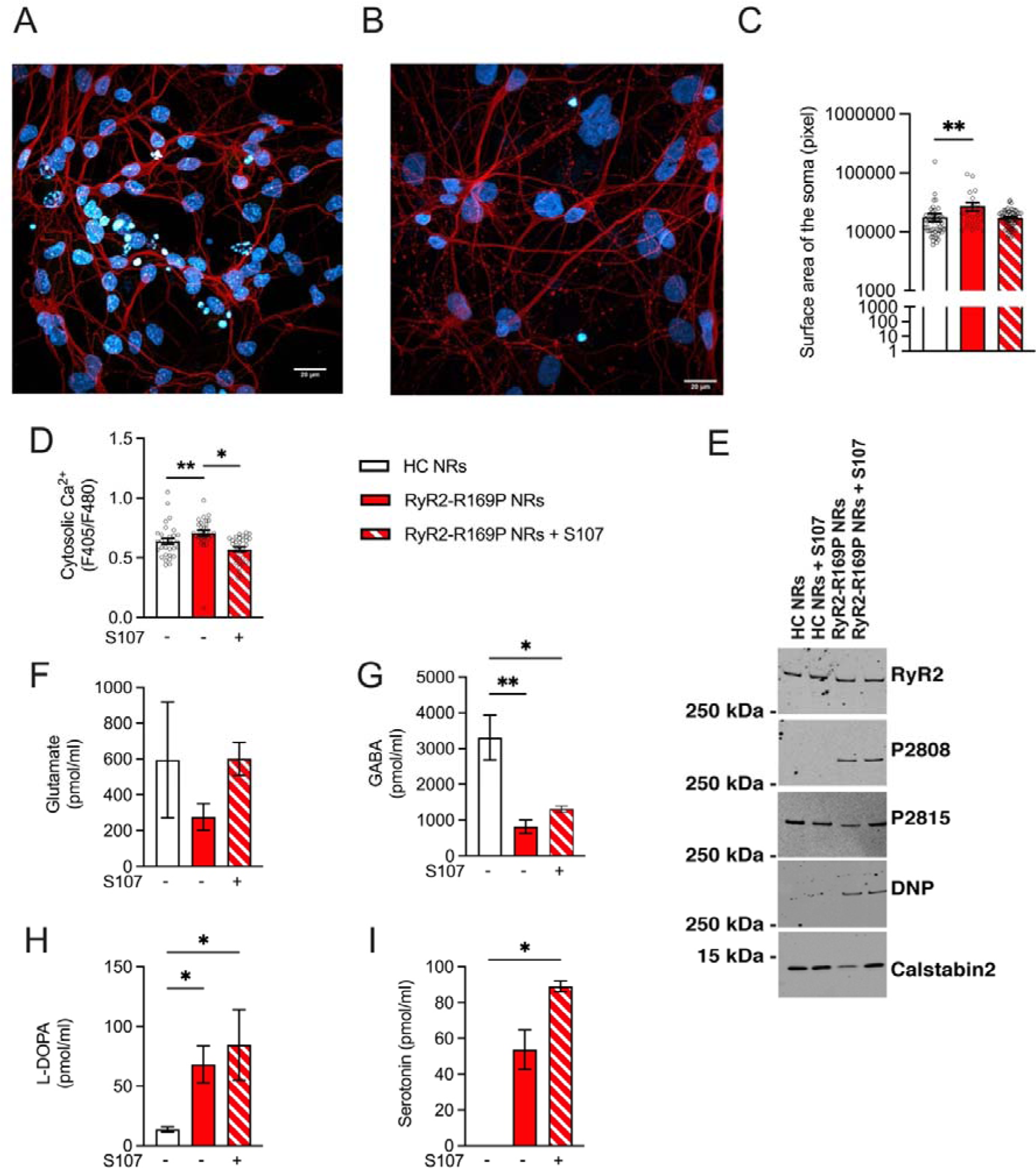
Neuronal characterization. Confocal immunofluorescence of HC (**A**) and RyR2-R169P (**B**) hiPSC-NRs stained with ß3-tubulin (red) and DAPI (blue). Scale bar: 20μm. **(C)** Scatter plot showing the surface of the soma in pixel in HC and RyR2-R169P hiPSC-NRs +/- S107. **(D)** Scatter plot showing the quantitative level of cytosolic Ca^2+^ in HC and RyR2-R169P hiPSC-NRs +/- S107 using 1µM Indo-1 AM dyes and IonOptix microscopy. **(E)** Immunoblots of RyR2 immunoprecipitated from protein extracts of HC and RyR2-R169P hiPSC-NRs without or with S107. P2808: Level of PKA phosphorylation at Serine 2809; P2815: Level of CaMKII phosphorylation at Serine 2815; DNP (2,4-dinitrophenylhydrazone): Level of RyR2 cysteine oxidation; Calstabin2: Level of calstabin2 depletion (for quantification see fig. S4). **(F, G, H and I)** Scatter plots showing the quantification of released glutamate, GABA, L-DOPA and serotonin neurotransmitters in HC and RyR2-R169P hiPSC-NRs +/- S107 (5μM overnight). Data are presented as mean ± SEM.

We further investigated the intracellular Ca^2+^ handling by quantitatively monitoring the Ca^2+^ levels in RyR2-R169P hiPSC-NRs with ratiometric Ca^2+^ indicator (Indo1-AM). We found elevated cytosolic Ca^2+^ levels in RyR2-R169P hiPSC-NRs, which was prevented by S107. These data suggested a link between intracellular Ca^2+^ leak and neuronal hypertrophy (Fig. 3D). Next, we monitored hiPSC-NPs at an early stage of neuronal differentiation (i.e., neuronal progenitors, NPs). We observed elevated cytosolic Ca^2+^ levels also in RyR2-R169P hiPSC-NPs, which was also prevented by S107 (Fig. 4A).

**Figure 4:**
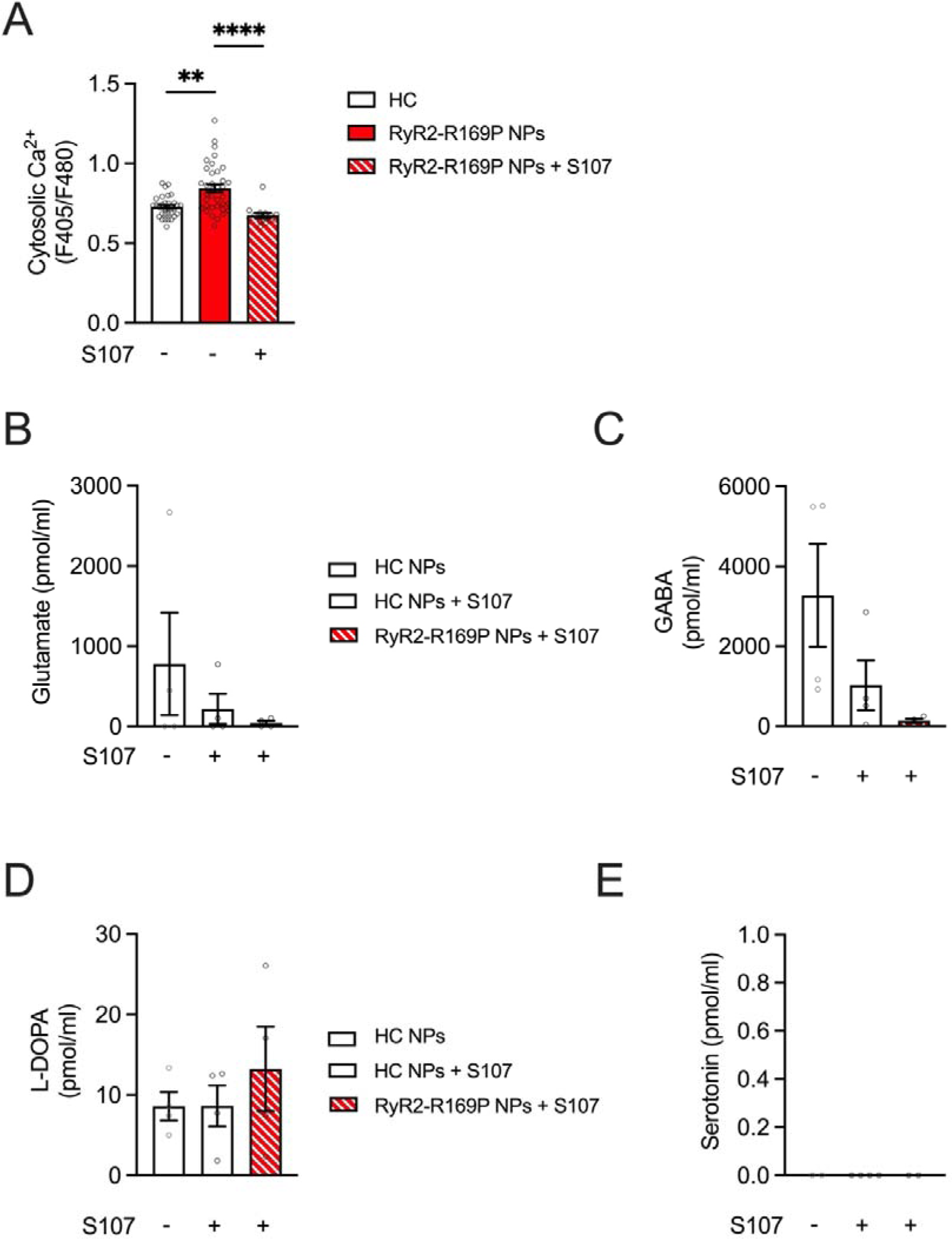
Effect of S107 on the NT release in RyR2-R169P hiPSC-NPs. **(A)** Scatter plot showing the quantitative level of cytosolic Ca^2+^ in HC and RyR2-R169P hiPSC-NPs +/- S107 using 1 µM Indo-1 AM dyes and IonOptix microscopy. **(B-E)** Scatter plots showing the quantification of released glutamate, GABA, L-DOPA and serotonin neurotransmitters in HC hiPSC-NRs +/- S107 and RyR2-R169P hiPSC-NRs + S107 (5μM for 6 weeks). Data are presented as mean ± SEM.

To test whether this neuronal defect is a feature common to all RyR2 mutations responsible for CPVT, we tested whether another RyR2 single-point mutations could lead to RyR2 leak in neurons and affect the neuronal cytosolic Ca^2+^ levels independently of the neuro-behavioral dysfunction observed in patients. We employed the RyR2-H29D hiPSC line and its corrected isogenic control from a patient suffering from ventricular tachycardia, who had no detected neuronal dysfunction ^16^. RyR2-H29D hiPSC-NRs did not exhibit elevated cytosolic Ca^2+^ levels compared to their isogenic control, regardless of the presence or absence of S107 (Fig. S3). All these data strongly suggest the specificity of the RyR2-R169P mutation in inducing RyR2 Ca^2+^ leak associated with neuronal dysfunction and ASD.

We then investigated whether the R169P mutation causes post-translational remodeling of the neuronal RyR2 macromolecular complex. Immunoblots of RyR2, immunoprecipitated from protein extracts of HC and RyR2-R169P hiPSC-NRs, were performed. We observed that the RyR2-R169P mutation results in higher PKA phosphorylation of RyR2 at Ser P2809, lower CaMKII phosphorylation of RyR2 at Ser P2815 and calstabin2 depletion with similar level of oxidized cysteines. When RyR2-R169P hiPSC-NRs were treated with S107, the level of RyR2 CaMKII phosphorylation was restored and calstabin2 depletion was prevented (Fig. 3E and S4).

### The RyR2-R169P mutation disrupts neurotransmitter homeostasis in midbrain hiPSC-NRs, mirroring neurochemical imbalances implicated in ASD

Mass spectrometry analysis revealed disruptions in neurotransmitter homeostasis in RyR2-R169P hiPSC-NRs. RyR2-R169P hiPSC-NRs showed a constant level of glutamate and a significant decrease in GABA release between RyR2-R169P and HC hiPSC-NRs (Fig. 3F-G) suggesting a disturbance of the excitation/inhibition (E/I) balance.

L-DOPA exhibited a significant increased level compared to HC cell lysate (Fig. 3H), indicative of dysregulated dopaminergic signaling. Additionally, an increased in serotonin secretion was also observed, further implicating perturbations in monoaminergic pathways. Notably, no serotonin was detected in HC cell lysates, underscoring the specificity of this alteration to the RyR2-R169P mutation (Fig. 3I). These findings align with the well-documented E/I imbalance in ASD, where elevated dopaminergic activity has been linked to behavioral phenotypes such as repetitive behaviors and social deficits ^18–20^. Importantly, treatment overnight of differentiated neurons with S107, failed to normalize neurotransmitter levels (Fig. 3F–G).

Given that basal calcium levels increase as early as the progenitor stage (Fig. 4A), even before neuronal differentiation, and considering that ASD is a neurodevelopmental disease, we hypothesized that normalizing basal calcium levels at the progenitor stage could prevent the changes described above. To test this hypothesis, we treated HC and RyR2-R169P hiPSC-NPs for 6 weeks during the differentiation stage. We found comparable levels of neurotransmitter release between HC +/- S107 and S107-treated RyR2-R169P hiPSC-NPs (Fig. 4B-E). These results highlight the importance of maintaining calcium homeostasis in neuronal differentiation and confirm the significance of RyR2 functional integrity in neuronal development.

### RyR2-R169P mutation results in abnormal teratoma neural crest development

Neurons and cardiomyocytes originate respectively from the ectodermal and mesodermal layers. We investigated whether the RyR2-R169P mutation affects neural development *in vivo*. To assess the mutation’s effect on *in vivo* neural development, we compared teratomas generated from HC and RyR2-R169P hiPSC lines in immunodeficient mice, with or without S107 treatment. While teratomas from both groups exhibited robust differentiation across all three germ layers (ectoderm, mesoderm, endoderm), RyR2-R169P teratomas displayed selective defects in neural crest-derived ectodermal structures. Specifically, mutant teratomas showed a significant reduction in dorsal root ganglion (DRG) neurons and a complete absence of melanocytes (Fig. 5A, D, E), critical cell types originating from the neural crest. In contrast, non-neural crest ectodermal derivatives, such as neural tube and choroid plexus structures, remained unaffected (Fig. 5A–C). Notably, S107 treatment failed to rescue these neural crest deficits, suggesting either inadequate drug bioavailability in teratomas or developmental-stage-specific irreversibility of the mutation’s effects. Mesodermal differentiation (into embryonic mesenchyme, cartilage, bone or smooth muscle) and endodermal differentiation (into lung or intestinal epithelium) were indistinguishable between groups (Fig. 5A, F–I), underscoring the mutation’s selective disruption of neural crest lineage maturation.

**Figure 5:**
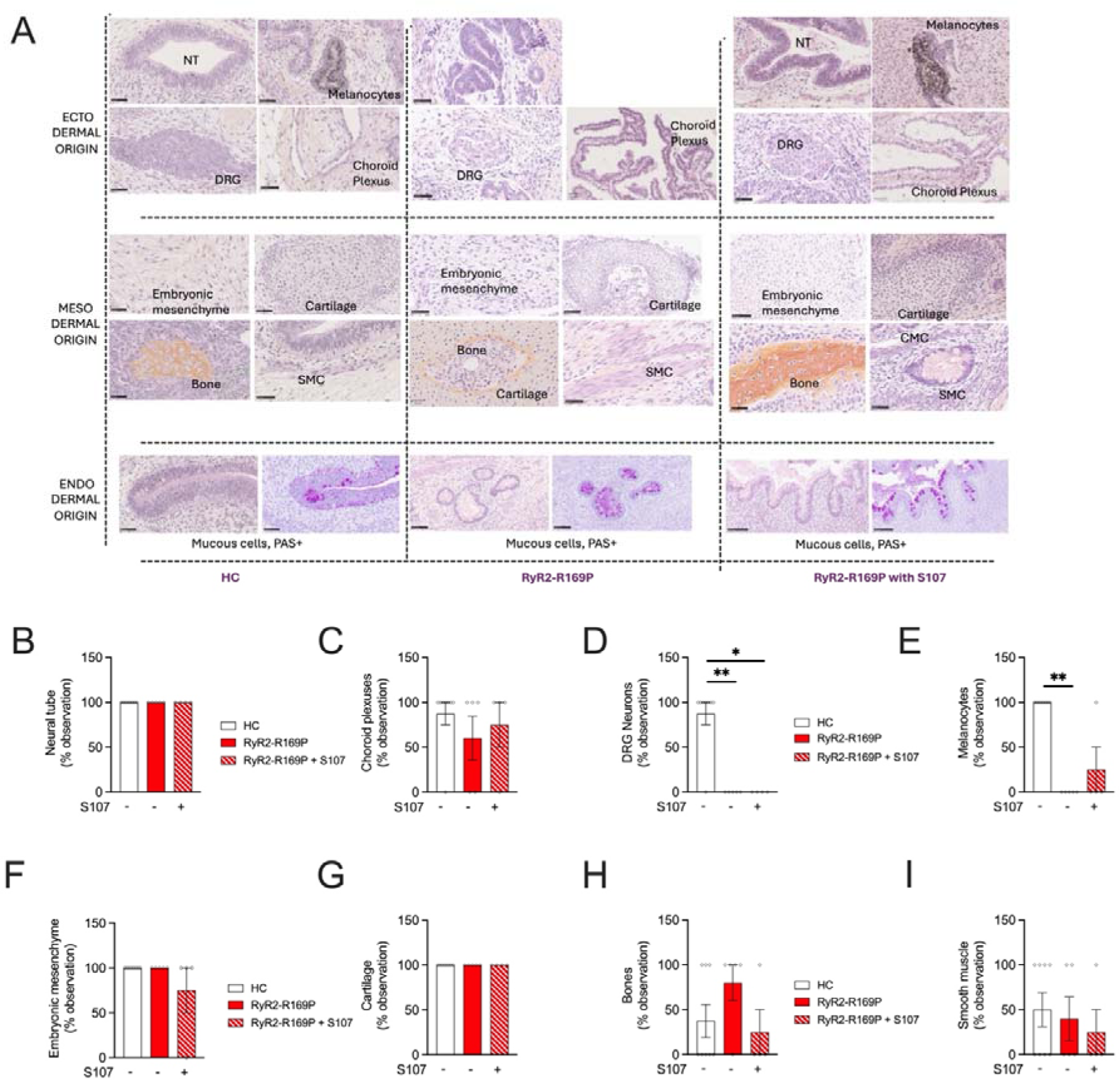
Characterization of the teratoma. **(A)** Analysis of the histological structures reveals cells and tissues derived from all three embryonic layers: ectoderm derivatives (neural tube (NT), melanocytes with their typical brown pigmentation, dorsal root ganglion (DRG) neurons and choroid plexus), mesoderm derivatives (embryonic mesenchyme, cartilage, bone and smooth muscle cells - SMC-) and endodermal derivatives (pulmonary and intestinal epithelium with mucinous cells characterized in the cytoplasm by fuschia-pink mucus droplets revealed by periodic acid-Schiff staining). **(B)** Scatter plots showing the percentage of observations of the neural tube from ectodermal layer. **(C)** Scatter plots showing the percentage of observations of the choroid plexus from ectodermal layer. **(D)** Percentage of observations of the DRG neurons from ectodermal layer. **(E)** Percentage of observations of the melanocytes from ectodermal layer. **(F)** Percentage of observations of the embryonic mesenchyme from mesodermal layer. **(G)** Percentage of observations of the cartilage from mesodermal layer. **(H)** Percentage of observations of the bones from mesodermal layer. **(I)** Percentage of observations of the smooth muscle from mesoderm. Data are presented as mean ± SEM.

### RyR2-R169P mutation leads to Ca^2+^ leak, calstabin2 depleted channels and abnormal intracellular calcium handling in CPVT hiPSC-CMs

We hypothesized that a pathological remodeling of the RyR2-R169P mutant channels leads to cardiac diastolic Ca^2+^ leak under stress. We conducted bilayer experiments to record RyR2 single-channel activity from HC and RyR2-R169P hiPSC-CMs previously exposed to ß-adrenergic stress (1µM). At a Ca^2+^ concentration of 150nM in the *cis* chamber (diastolic condition), we observed an increased single-channel open probability (Po) of the PKA-phosphorylated RyR2-R169P channels (Fig. S5A, B and C). This elevated Po was accompanied by a higher frequency of channel openings, while the RyR2 mean closed and open times remained unaffected (Fig. S5A, B, D, E and F).

We further investigated whether the novel RyR2-R169P mutation leads to an abnormal intracellular cycling in hiPSC-CMs by measuring the Ca^2+^ dynamics under stress condition using isoproterenol (1µM). We found that RyR2-R169P hiPSC-CMs exhibited alternating Ca^2+^ transients, a pathological pattern that was prevented by S107 (Fig. 6A). Compared to HC hiPSC-CMs, RyR2-R169P hiPSC-CMs showed reduced amplitude and velocity (Ca^2+^ release time indicator), and similar Ca^2+^ transient frequency (Fig. 6A, B, D and E). The decay time (Ca^2+^ reuptake time indicator) was similar in all conditions (Fig. 6C). S107 treatment was able to prevent the reduction in amplitude and velocity and also reduced the Ca^2+^ transient frequency compared to HC hiPSC-CMs. Additionally, we quantitatively measured the diastolic Ca^2+^ level and found that RyR2-R169P hiPSC-CMs exhibited higher Ca^2+^ levels under stress conditions, which were reduced to levels comparable to those of HC hiPSC-CMs by S107 treatment (Fig. 6F).

**Figure 6:**
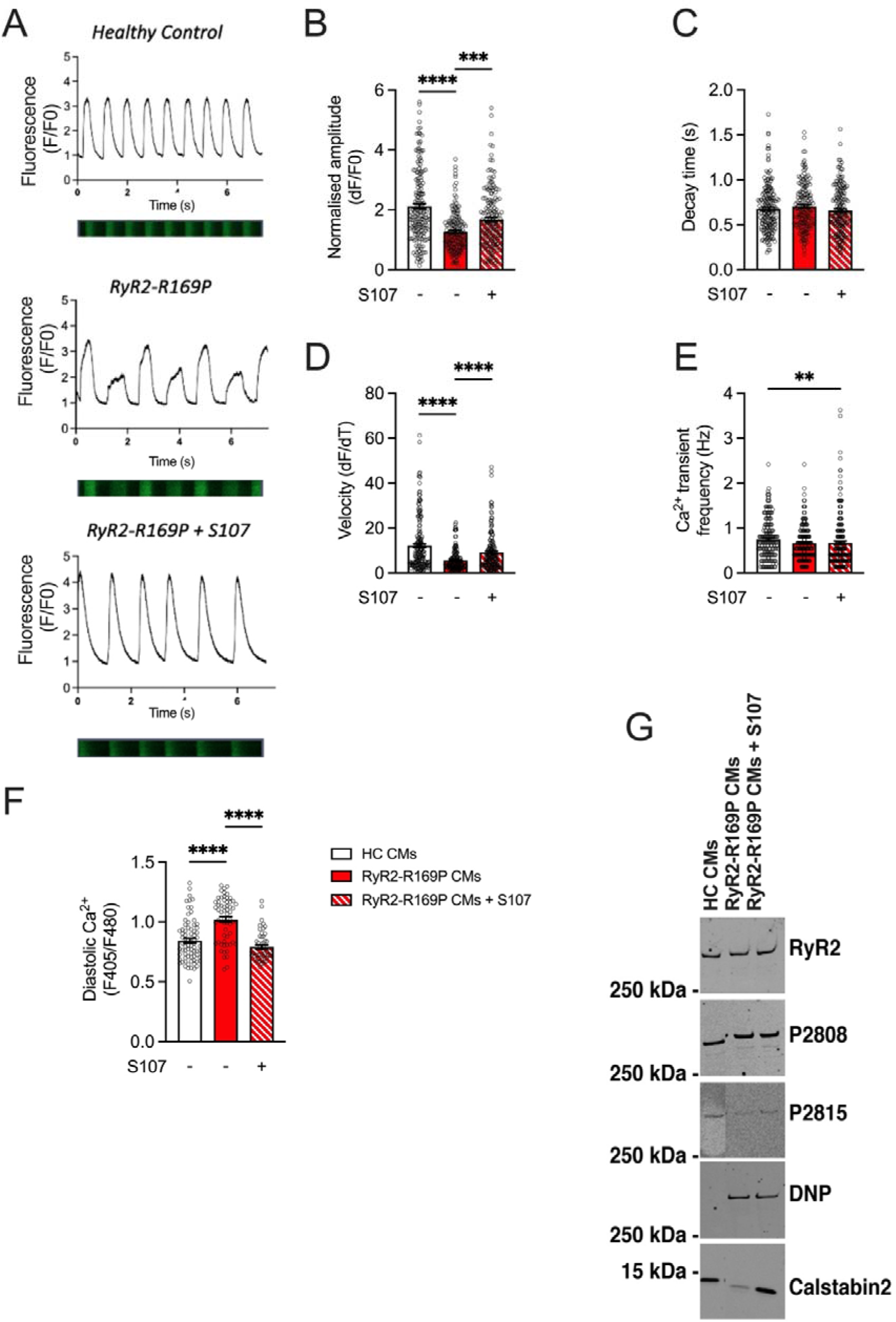
Characterization of the intracellular Ca^2+^ cycling in HC and RyR2-R169P hiPSC-CMs. **(A)** Original line-scan image of Ca^2+^ transients and corresponding tracing in HC and RyR2-R169P hiPSC-CMs +/- S107. **(B)** Scatter plot showing the normalized amplitude of the Ca^2+^ transients in HC and RyR2-R169P hiPSC-CMs +/- S107. **(C)** Scatter plot showing the decay time (s) in HC and RyR2-R169P hiPSC-CMs +/- S107. **(D)** Velocity (dF/dT) in HC and RyR2-R169P hiPSC-CMs +/- S107. **(E)** Ca^2+^ transient frequency (Hz) in HC and RyR2-R169P hiPSC-CMs +/- S107. **(F)** Scatter plot showing the quantitative level of diastolic Ca^2+^ in HC and RyR2-R169P hiPSC-CMs +/- S107. **(G)** Immunoblots of RyR2 immunoprecipitated from protein extracts of HC and RyR2-R169P hiPSC-CMs without or with S107. P2808: Level of PKA phosphorylation at Serine 2809; P2815: Level of CaMKII phosphorylation at Serine 2815; DNP (2,4-dinitrophenylhydrazone): Level of RyR2 cysteine oxidation; Calstabin2: Level of calstabin2 depletion. Data are presented as mean ± SEM.

We have previously reported that mutated RyR2 channels exhibit pathological remodeling in hiPSC-CMs ^4, 5^. We tested whether RyR2-R169P causes post-translational remodeling under ß-adrenergic stress conditions. Immunoblots of RyR2, immunoprecipitated from protein extracts of HC and RyR2-R169P hiPSC-CMs, were performed. We observed that the RyR2-R169P mutation results in calstabin2 depletion of the RyR2 macromolecular complex (Fig. 6G and S6D). When RyR2-R169P hiPSC-CMs were treated with S107, calstabin2 depletion was prevented (Fig. 6G and S6D). The PKA and CaMKII phosphorylation levels were unchanged between HC and RyR2-R169P hiPSC-CMs (Fig. 6G and S6A, B and C).

We evaluated the impact of the RyR2-R169P mutation on the cell morphology of the hiPSC-CMs by immunofluorescence. Cell area and sarcomere length were unaffected by the mutation (Fig. S7).

### The RyR2-R169P mutation would destabilize the local structure of RyR2, directly linking its 3D conformational defects to Ca^2+^ leak pathology

The R169 residue has previously been associated with CPVT syndrome through two other single-point mutations, RyR2-R169Q and RyR2-R169L ^21–23^. *In silico* structural modeling of the novel RYR2c.506G>C mutation (resulting in RyR2-R169P) revealed its localization within a rigid loop of the N-terminal domain-A (NTD-A), a region stabilized by a critical hydrogen bond network involving residues R169, E173, R176, and D179 (Fig. S8A–D). Interestingly, the residue R176 in this hydrogen bond network has also been associated with CPVT with the RyR2-R176Q mutation, and the structure resolved by cryo-EM ^24^. Disruption of this hydrogen bond network as seen by cryo-EM leads to rotation of the NTD-A domain as seen in the primed state ^24^. This mechanism of local destabilization and rotation of the NTD domain, which leads to the pathological leaky primed state, has also been reported by cryo-EM for other CPVT mutations (ref https://www.nature.com/articles/s41467-024-51791-y). Comparative analysis of the three CPVT-associated mutations at position 169 (R169Q, R169L, R169P) demonstrated escalating disruption: i/ RyR2-R169Q would partially destabilize hydrogen bonds, mirroring the conformational “primed state” seen in the RyR2-R176Q mutant (Fig. S8F), which weakens interactions with adjacent NTD-B* and CSol* domains. ii/ RyR2-R169L would cause greater hydrogen bond disruption due to its hydrophobic nature, correlating with more severe clinical phenotypes. iii/ RyR2-R169P would exhibit the most profound structural impact, destabilizing both hydrogen bonding and backbone geometry (Fig. S8G–I), rendering the channel hyperactive under stress.

In RyR2-WT, Ca^2+^ binding triggers coordinated pore opening and a downward-outward shift of the cytoplasmic shell (high energy cost). CPVT mutants like R169P lock the cytoplasmic shell in an intermediate “primed” conformation (Fig. S8C–E), decreasing the energy barrier and promoting pathological Ca^2+^ leak. Critically, the RyR2 stabilizer S107 reversed this defect by restoring an upward-inward cytoplasmic shift, resealing the channel (Fig.S87J). These structural insights explain the mutation’s dual cardiac and neuronal impact, as destabilized RyR2 primes cells for Ca^2+^-driven dysfunction under stress.

## Discussion

CPVT, a rare inherited rhythmic disorder (1/10000-1/15000), is often associated with neurobehavioral disorders. Our study focuses on the ASD comorbidity associated with 15-20% of patients with CPVT. The clinical characterization of our patient, who was diagnosed with both ASD and CPVT, revealed no signs of cerebral suffering *in utero*, neonatal periods, or during a pool accident, ruling out hypoxic-ischemic encephalopathy as a contributing factor. The patient did not experience epileptic seizures. He exhibited neurodevelopmental disorders, including dysphasia and dyspraxia, with normal genetic, metabolic, and brain imaging analyses. The diagnosis of CPVT was confirmed after an episode of bidirectional ventricular tachycardia, accompanied by the identification of the novel RyR2-R169P mutation. The mosaicism of RyR2-R169P in the asymptomatic mother suggests variable penetrance or compensatory mechanisms, warranting further study into genetic modifiers.

We employed the patient-specific RyR2-R169P hiPSC line previously characterized ^13^ to investigate the impact of this novel RyR2 mutation on hiPSC-CMs and -NRs. As previously demonstrated ^13^, we conducted a bulk reprogramming to generate a large number of hiPSCs simultaneously, enhancing consistency and reducing variability. Our objective was to utilize CRISPR/Cas9 technology to correct the single-point mutation RyR2-R169P in hiPSCs and study the genotype/phenotype relationship using the latest cutting-edge technology. We implemented two distinct strategies to achieve this correction. However, both approaches were unsuccessful, likely due to the inaccessibility of the target DNA site within the chromatin structure. This failure highlights the challenges of targeting certain regions of the genome for precise genetic modifications, a challenge increasingly recognized in genomic regions with repressive epigenetic marks or repetitive sequences ^25^. Consequently, we employed a male HC hiPSC line previously characterized ^11^. Midbrain neurons are affected in ASD, exhibiting abnormal morphology and function, especially those involved in dopaminergic pathway ^26^. Our results revealed that the midbrain hiPSC-NRs harboring RyR2-R169P mutation exhibit abnormal intracellular Ca^2+^ homeostasis with an increased basal Ca^2+^ level that is prevented by S107 specifically targeting RyR2 as we previously demonstrated ^4, 27^.

The study of intracellular neuronal Ca^2+^ dynamics revealed a disturbance in NRs, indicating the involvement of RyR2. These findings are consistent with those observed at the cardiac level. However, treatment with S107 did not completely restore these parameters. If S107 failed to prevent these alterations in intracellular Ca^2+^ dynamics, it may be due to neuronal Ca^2+^ kinetics involving both RyR2 and, to a greater extent, IP_3_R. This observation may also be attributed to the fact that ASD is a neurodevelopmental disorder, suggesting that S107 treatment should be administered to NRs at an earlier stage and during differentiation. Furthermore, we found that cytosolic Ca^2+^ levels are more abundant in RyR2-R169P adult and progenitor hiPSC-NRs compared to HC counterparts. Our results indicated that the RyR2-R169P mutation causes an increase in cytosolic Ca^2+^ beginning at the early progenitor stage, potentially corresponding to developmental periods when the first signs of ASD may emerge. These increases involve a RyR2 Ca^2+^ leak from the endoplasmic reticulum to the cytosol. Interestingly, S107 corrects the Ca^2+^ leak at both the neuronal and progenitor levels. Our results suggest, for the first time, a link between a RyR2 mutation and an ASD phenotype.

To better understand the impact of the RyR2-R169P mutation on CPVT syndrome associated with ASD, we investigated whether other RyR2 mutations could induce similar Ca^2^□ disturbances potentially leading to undiagnosed ASD. We utilized a hiPSC line harboring the RyR2-H29D mutation from a patient with polymorphic ventricular tachycardia ^5^. This mutation, also located in the N-terminal part of RyR2, provided a relevant comparison. The RyR2-H29D hiPSC line had the advantage of an isogenic corrected control created using CRISPR/Cas9 technology. Measurements of cytosolic Ca^2^□ levels in both RyR2-H29D hiPSC-NRs and their isogenic controls showed no significant differences. These results suggest that not all RyR2 mutations lead to alterations in Ca^2^□ homeostasis, indicating that the ASD observed in the patient with the RyR2-R169P mutation may be specifically linked to this particular mutation.

While the RyR2-R169P mutation induces Ca^2^□ dysregulation and neurodevelopmental abnormalities (*e.g.*, neuronal hypertrophy, neurotransmitter imbalances), the absence of seizures and normal EEG results suggest that the mutation does not directly perturb neuronal hyperexcitability or synchronized network activity to a pathological extent. Consequently, we did not perform electrophysiological recordings with the RyR2-R169P hiPSC-NRs. This contrasts with other RyR2 mutations, such as the RyR2-R2474S mutation, which has been associated with epilepsy ^6^. The data reinforce that the observed ASD-related phenotypes in this patient arise from Ca^2+^-dependent developmental or metabolic disruptions rather than primary electrophysiological instability in the CNS.

To further confirm the impact of the RyR2-R169P mutation on neural development, we generated teratomas in mice to assess the differentiation potential of hiPSCs into various cell types derived from the three germ layers, providing a comprehensive model to study the effects of the RyR2-R169P mutation *in vivo*. Our observations revealed altered differentiation in neural crest-derived ectodermal cells in RyR2-R169P teratomas, characterized by a reduced number of DRG neurons and an absence of melanocytes. While speculative, RyR2 Ca^2+^ leak during early neurogenesis could disrupt neural crest cell commitment, migration or differentiation, potentially affecting brain regions implicated in ASD, such as the anterior cingulate cortex or striatum. Additionally, the enlarged soma size observed in RyR2-R169P neurons suggests aberrant neuronal growth, consistent with findings from a hiPSC model of ASD reported by Marchetto *et al.* ^28^. It may compromise circuit formation during critical developmental windows. Though not directly assessed here, RyR2 dysfunction is linked to mitochondrial Ca^2+^ overload and oxidative stress in other models as already reported for other RyR2-related pathologies ^10^. These pathways could exacerbate neuronal vulnerability, particularly in energy-demanding processes like synaptic remodeling, further contributing to ASD-related phenotypes.

Notably, S107 treatment did not prevent these alterations in the ectodermal layer, specifically in neural crest differentiation. This lack of preventive effect may be attributed to the fact that S107 administered in drinking water likely did not reach the subcutaneous teratomas effectively. The decrease in cell numbers may be attributed to either a reduction in the production of ectodermal cells or their derivatives, or an increase in the mortality of ectoderm-derived cells. This approach underscores the importance of using teratomas to evaluate the broader developmental impact of RyR2 mutations.

Functional studies of RyR2-R169P hiPSC-CMs were conducted with a primary focus on Ca^2+^ dynamics involved in cardiac excitation-contraction coupling under β-adrenergic receptor stress, a condition during which CPVT manifests. Our results demonstrated that the RyR2-R169P mutation causes aberrant SR Ca^2+^ cycling with elevated diastolic Ca^2+^ level under β-adrenergic receptor stress as previously reported for several other RyR2 mutations ^4^. We observed an increased Po in RyR2-R169P under stress at resting Ca^2+^concentrations, suggesting that this mutation confers a gain-of-function to the mutant channels. Our results also evidenced the pathological remodeling of the RyR2 macromolecular complex under stress. Lower CaMKII phosphorylation at S2815, increased oxidized cysteines and calstabin2 depletion were found while the PKA phosphorylation at S2809 was unaffected. Treatment of RyR2-R169P hiPSC-CMs with S107 successfully prevented the depletion of calstabin2 and prevented the aberrant SR Ca^2+^ cycling with normal diastolic Ca^2+^ level. RyR2-R169P hiPSC-CMs did not exhibit any morphological alteration in sarcomere size, in line with clinical observations and the literature regarding the structure of CPVT patient cardiomyocytes ^29, 30^.

To get insight into the neuro-behavioral dysfunction clinically revealed in this patient, we investigated patient-specific midbrain hiPSC-NRs. We investigated the morphology and showed that the RyR2-R169P mutation leads to a modification of the soma size while the nuclei are normal. S107 treatment reversed the surface area of patient neurons’ somas to normal levels.

Our study reveals that the RyR2-R169P mutation induces pathological remodeling of the RyR2 macromolecular complex in hiPSC-NRs, characterized by reduced CaMKII phosphorylation at Ser2815 and calstabin2 depletion. This hypo-phosphorylation was rescued by S107, which also restored calstabin2 binding and normalized cytosolic Ca^2^□ levels. These findings align with emerging evidence implicating CaMKII dysregulation in neurodevelopmental disorders, including ASD. CaMKII is a critical mediator of synaptic plasticity, and its activity modulates key synaptic proteins such as Shank3, a scaffolding protein genetically linked to ASD ^31^. Phosphorylation of Shank3 by CaMKII regulates synaptic strength and dendritic spine morphology ^32^, and mutations in Shank3 disrupt this interaction, contributing to ASD-associated synaptic deficits ^31^. Notably, a CaMKIIα mutation (E183V) identified in ASD patients alters synaptic transmission and dendritic architecture, recapitulating ASD-like behaviors in mice ^32^. Our observation that RyR2-R169P neurons exhibit reduced CaMKII phosphorylation suggests a novel mechanism by which RyR2 dysfunction may perturb CaMKII-dependent synaptic signaling. The restoration of CaMKII phosphorylation by S107 further supports the therapeutic potential of targeting RyR2 stabilization to mitigate Ca^2+^-driven synaptic anomalies in ASD.

Ca^2+^ signaling through RyR2 is essential for activity-dependent synaptic plasticity, including long-term potentiation (LTP) and depression (LTD). Our data show that RyR2-R169P neurons exhibit hypo-phosphorylation of CaMKII at Ser2815, a kinase critical for stabilizing synaptic strength and dendritic spine morphology via targets like Shank3. Disrupted CaMKII-Shank3 interactions, as observed in ASD models, could destabilize excitatory synapses and impair the refinement of neural circuits during development. These synaptic defects may underlie the impaired social learning and communication skills characteristic of ASD. Further experiments are needed to investigate the downstream effects on synaptic plasticity proteins, such as Shank3 and PSD-95, to elucidate the mechanistic link to ASD.

We observed significant alterations in neurotransmitter levels in RyR2-R169P neurons, in particular an increased L-DOPA and serotonin release and reduced GABA levels. These changes are relevant to ASD, as neurotransmitter imbalances are known to contribute to the behavioral and cognitive symptoms of the disorder. The decreased GABA levels suggest an excitatory/inhibitory imbalance. Reduced GABAergic tone, as seen in RyR2-R169P neurons, could impair inhibitory synaptic transmission, potentially leading to network hyperexcitability and disrupted information processing in brain regions such as the prefrontal cortex and amygdala. The elevated L-DOPA level indicate dysregulation of dopaminergic pathway, a pathway implicated in repetitive behaviors. These neurotransmitters alterations, driven by RyR2-mediated Ca^2+^ leak, provide a mechanistic bridge between genetic mutation and behavioral phenotypes. Our findings provide insights into the neurochemical basis of ASD in the context of RyR2 mutations. They are in line with previous reports showing that dopamine, serotonin, and the excitatory/inhibitory imbalance leads to disruptions in neurodevelopmental anomalies of the CNS that can lead to ASD ^33, 34^.

The inability of S107 to restore neurotransmitter levels when applied acutely to mature neurons demonstrates that the RyR defect is not directly involved in the devaluation of neurotransmitter release. In contrast, the ability of S107 to normalize neurotransmitter release when applied chronically from the progenitor stage demonstrates and confirms the importance of RyRs in neurodevelopmental processes and in neuronal differentiation leading to a normal releasing profile.

Our results suggest two hypotheses that could explain the impact of the RyR2-R169P mutation at the neuronal level. Firstly, the location of the mutation and the specific amino acids involved are crucial. The mutation in the patient with CPVT and ASD is located in the N-terminal part of RyR2, a region sensitive to mutations linked to CPVT, at position 169. This mutation involves the substitution of a positively charged amino acid arginine with a neutral proline. Under physiological conditions, the arginine at position 169 stabilizes the N-terminal domain by interacting with residues R176 and D179, forming a stabilizing loop. In the pathological context of the RyR2-R169P mutation, the substitution of arginine with proline likely and the lack of positive charges disrupts electrostatic interactions, destabilizing the loop and leading to a primed state of the channel. Secondly, there may be an alteration in the interaction between RyR2 and the L-type Ca^2+^ channel in neurons.

Despite the promising findings, our study has several limitations. The immaturity of hiPSC-derived cells may not perfectly recapitulate the adult phenotype, potentially affecting the translatability of the results. Our hiPSC-derived neuronal models may not fully recapitulate the complexity of *in vivo* neural networks or account for non-cell-autonomous factors (*e.g.,* glial interactions, immune signaling). The absence of behavioral data or direct evidence linking Ca^2+^ dysregulation to social/communication deficits in this patient limits causal inference. Additionally, the lack of an isogenic control limits the ability to attribute observed phenotypes solely to the RyR2-R169P mutation. However, while the absence of an isogenic control limits causal attribution to RyR2-R169P, the specificity of the intracellular Ca^2+^ defects, absent in RyR2-H29D neurons, strengthens our conclusions. Future studies should address these limitations by using more mature cell models and generating isogenic controls when possible. The use of Fluo-4/INDO-1 dyes may not capture subtle intracellular Ca^2+^ dynamics. The contribution of IP_3_R vs. RyR2 to neuronal Ca^2+^ transients is not dissected, complicating interpretation.

Future research should explore mitochondrial and inflammatory pathways and validate findings in vivo. This study provides comprehensive insights into the molecular mechanisms underlying CPVT and ASD comorbidity, highlighting potential therapeutic strategies for managing these disorders.

## Conclusion

Our study establishes RyR2-R169P as a dual regulator of cardiac and neuronal Ca^2+^ homeostasis, offering a unified mechanism for CPVT-ASD comorbidity. The efficacy of S107 in rescuing Ca^2+^ handling defects underscores its potential as a targeted therapy, while patient-derived hiPSC models pave the way for personalized treatment strategies.

RyR dysfunction is not exclusively associated with point mutations in the channel. Indeed, we have previously described central nervous system pathologies such as PTSD ^8^, Alzheimer’s disease ^10^, Huntignton’s disease ^35^ or cardiogenic dementia ^36^ associated with post-translational modifications of RyR2 responsible for cytosolic calcium leakage. These observations allow us to suggest that *in-utero* situations of antenatal stress responsible for early post-translational modifications of RyR2 during neurodevelopment could explain the appearance of sporadic forms of autism spectrum disorder associated with no mutation.

## Contributors

SC conducted the experiments on the hiPSC-CMs and performed the statistical analyses. SC also carried out the experiments on cardiac and neuronal differentiation, the experiments on ICC, Ca^2+^ imaging and contractility. SR conducted experiments on biochemistry. SC, HB, and AAB conducted statistical analyses. MM performed the 3D *in silico* protein modeling. JLP provided clinical data and medical expertise. SC ACM and AL wrote the manuscript. JLP, ARM, AL, and ACM designed the study, interpreted the data, and provided edits to the manuscript. FB performed histopathological analysis. All authors critically reviewed the manuscript and approved its submission.

## Declaration of Interests

ARM is a board member and owns shares in RyCarma Therapeutics, which is targeting RyR channels for therapeutic purposes. All authors have nothing else to disclose. All authors read and approve the final version of the manuscript, and ensure it is the case.

## Funding

This work was supported by grants of the “Institut National pour la Santé et la Recherche Médicale” (INSERM), ANR Musage and Fenice and the “Fondation Coeur et Recherche” of the French Society of Cardiology, the “Fondation pour la Recherche Médicale” and GRRC (Thesis grant for SC). The funders had no role in study design, data of collection, data analysis, interpretation or writing of the report.

SMPMS work was funded by INSERM, CNRS, University of Strasbourg (Unistra and CoRTecS; DIALOG 2024 Ref.: RBA/SCC/JLA/DHA/N° 2023-464m and RBA/SCC/JLA/DHA/N° 2023-464m), the Interdisciplinary Thematic Institute NeuroStra, as part of the ITI 2021-2028 program of the University of Strasbourg, CNRS and lnserm, was supported by ldEx Unistra (ANR-10-IDEX-0002) under the framework of the French Program Investments for the Future. SMPMS was also funded by the Agence Nationale de la Recherche (ANR) under the project “ANR-22-CE18-0037-03” the FRC Neurodon – Rotary – “Espoir en Tête 2022”.

## Data Availability

All data produced in the present study are available upon reasonable request to the authors.

## Acknowledgements

We are grateful for the technical support of the RHEM (Réseau d’Histologie Expérimentale de Montpellier) histopathology facility.

---

**Table S1:** LC-MS/MS conditions. LC and MS/MS conditions for the purification, detection and quantification of dopamine, serotonin, noradrenaline, GABA and glutamate their respective heavy-tagged counterparts. The flow rate was set at 90 µl/min on a ZORBAX SB-C18 column (150 x 1mm, 3.5μm).

|  | Mobile phase |  |  |
| --- | --- | --- | --- |
|  | ACN | H<sub>2</sub>O | Formic acid |
| Mobile phase A | 1% | 98.9% | 0.1% |
| Mobile phase B | 99.9% | 0 | 0.1% |

| HPLC gradient |  |  |  |  |  |  |  |
| --- | --- | --- | --- | --- | --- | --- | --- |
| Time (min) | 0 | 2.5 | 9 | 11 | 12 | 12.5 | 18 |
| % B mobile phase | 0 | 0 | 20 | 98 | 98 | 0 | 0 |

| MS parameters |  |
| --- | --- |
| Mode | positive |
| Spray voltage | 3,500 V |
| Nebulizer gas | Nitrogen |
| Desolvation (nitrogen) sheath gas | 18 Arb |
| Aux gas | 7 Arb |
| Ion transfer tube temperature | 297°C |
| Vaporizer temperature | 131°C |
| Q1 and Q3 resolutions | 0.7 FWHM |
| Collision gas (CID, argon) pressure | 2 mTorr |

| Compound | Polarity | Precursor (m/z) | Product (m/z) | Ion product type | Collision Energy (V) | RF Lens (V) |
| --- | --- | --- | --- | --- | --- | --- |
| Dopamine | Positive | 324.14 | 116.11 | Quantification | 54.24 | 206 |
| Dopamine | Positive | 324.14 | 145.11 | Qualification | 33.36 | 206 |
| Dopamine | Positive | 324.14 | 171.06 | Qualification | 23.90 | 206 |
| D4-Dopamine | Positive | 328.14 | 122.11 | Quantification | 35.00 | 206 |
| Noradrenaline | Positive | 340.15 | 116.11 | Quantification | 55.00 | 215 |
| Noradrenaline | Positive | 340.15 | 145.11 | Qualification | 36.19 | 215 |
| Noradrenaline | Positive | 340.15 | 171.06 | Qualification | 24.61 | 215 |
| C6-Noradrenaline | Positive | 346.15 | 145.11 | Quantification | 36.04 | 226 |
| C6-Noradrenaline | Positive | 346.15 | 158.11 | Qualification | 18.34 | 226 |
| C6-Noradrenaline | Positive | 346.15 | 171.06 | Qualification | 25.62 | 226 |
| GABA | Positive | 274.11 | 116.04 | Quantification | 46.35 | 157 |
| GABA | Positive | 274.11 | 128.11 | Qualification | 43.47 | 157 |
| GABA | Positive | 274.11 | 171.06 | Qualification | 19.66 | 157 |
| D6-GABA | Positive | 280.16 | 116.11 | Quantification | 48.73 | 150 |
| D6-GABA | Positive | 280.16 | 128.04 | Qualification | 44.78 | 232 |
| D6-GABA | Positive | 280.16 | 171 | Qualification | 19.60 | 232 |
| Glutamate | Positive | 318.11 | 116.11 | Quantification | 55.00 | 169 |
| Glutamate | Positive | 318.11 | 128.11 | Qualification | 52.82 | 169 |
| Glutamate | Positive | 318.11 | 170.93 | Qualification | 20.52 | 169 |
| D5-Glutamate | Positive | 323.12 | 116.11 | Quantification | 55 | 159 |
| D5-Glutamate | Positive | 323.12 | 128.06 | Qualification | 48.68 | 159 |
| D5-Glutamate | Positive | 323.12 | 171.00 | Qualification | 20.77 | 159 |

## Notes

### Funding Statement

This work was supported by grants of INSERM, ANR Musage and Fenice and the Fondation Coeur et Recherche of the French Society of Cardiology.

### Author Declarations

This study was conducted in accordance with the Declaration of Helsinki and was approved by the Montpellier Hospital Review Board Committee (approval number 1003-HPS2).

