## Supplemental figures for "Patient-derived cells demonstrate that mutated RyR2 calcium leak underlies autism spectrum disorder and inherited arrhythmias"

**Supplementary figures**

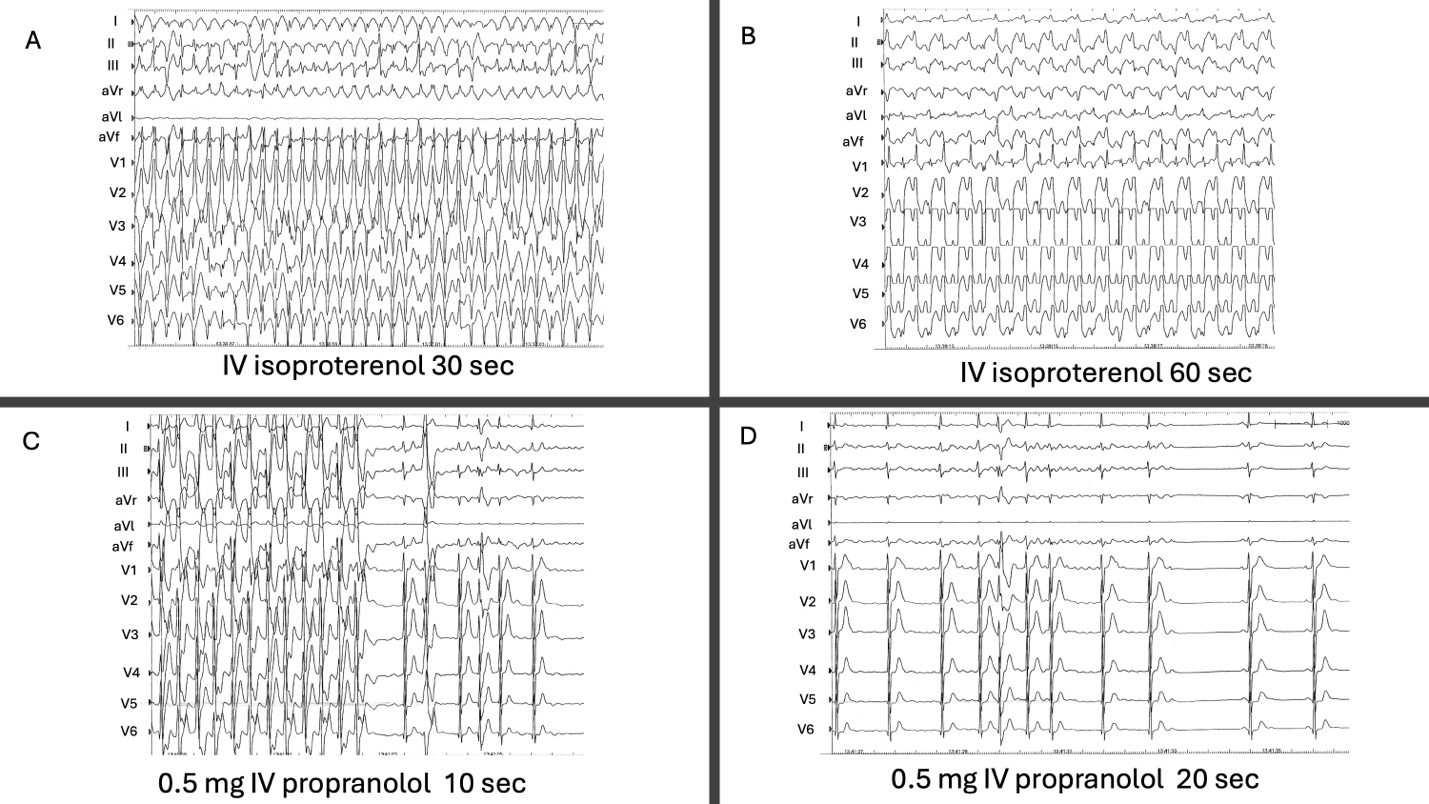

**Figure S1: (A)** Pharmacological testing: Intravenous infusion of isoproterenol increases heart rate, and within 30 seconds, rapid atrial fibrillation is observed. **(B)** After 60 seconds isoproterenol infusion induces bidirectional ventricular tachycardia, characteristic of CPVT. **(C)** Following the induction of bidirectional VT, isoproterenol was stopped, and IV propranolol (0.5 mg) was administered. Within 10 seconds, bidirectional VT ceased, revealing atrial fibrillation. **(D)** Within 20 seconds after IV propranolol administration, AF stopped and converted to sinus rhythm.

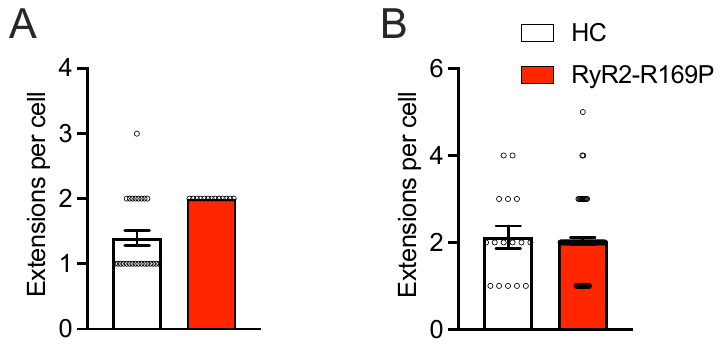

**Figure S2: (A)** Scatter plot showing the number of extensions per neuron at the progenitor state in HC and RyR2-R169P hiPSC-NPs. **(B)** Scatter plot showing the number of extensions per neuron at the adult state in HC and RyR2-R169P hiPSC-NRs. Data are presented as mean ± SEM.

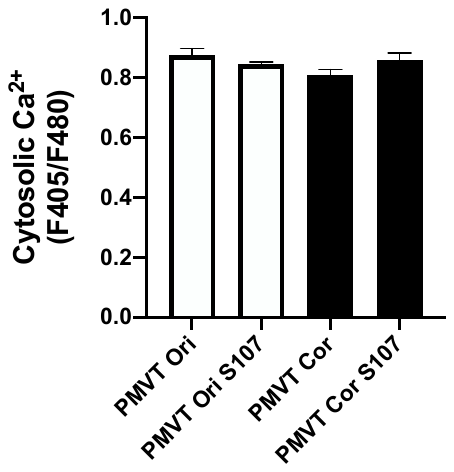

**Figure S3:** Scatter plot showing the quantitative level of cytosolic Ca^2+^ in RyR2-H29D (Ori) and isogenic control (Cor) hiPSC-NRs +/- S107. Data are presented as mean ± SEM.

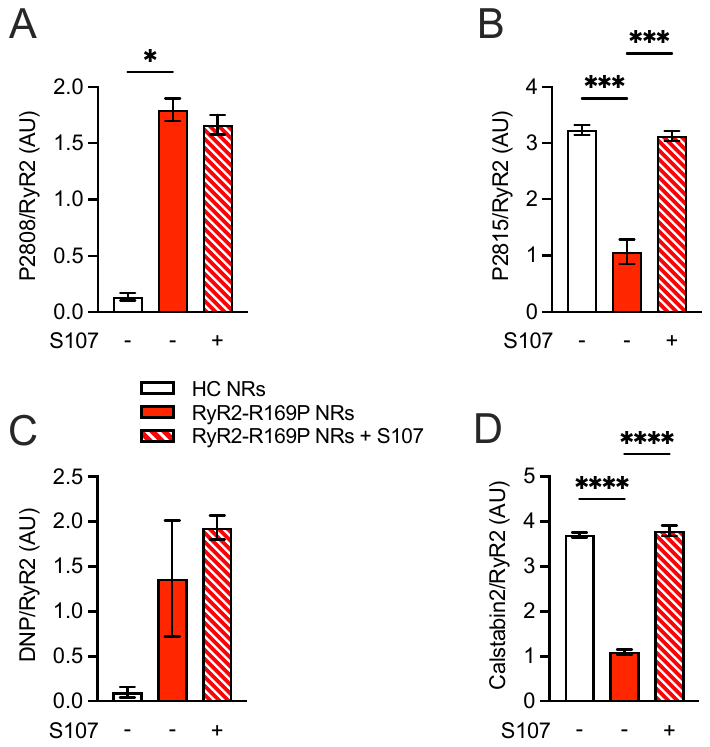

**Figure S4: (A)** Bar graphs showing the relative RyR2 PKA-phosphorylation level at Ser 2809 in HC and RyR2-R169P hiPSC-NRs +/- S107. **(B)** Relative RyR2 CaMKII-phosphorylation level at Ser 2815 in HC and RyR2-R169P hiPSC-NRs +/- S107. **(C)** Relative RyR2 oxidation level in HC and RyR2-R169P hiPSC-NRs +/- S107. **(D)** Relative calstabin2 amount bound to RyR2 in HC and RyR2-R169P hiPSC-NRs +/- S107. Data are presented as mean ± SEM.

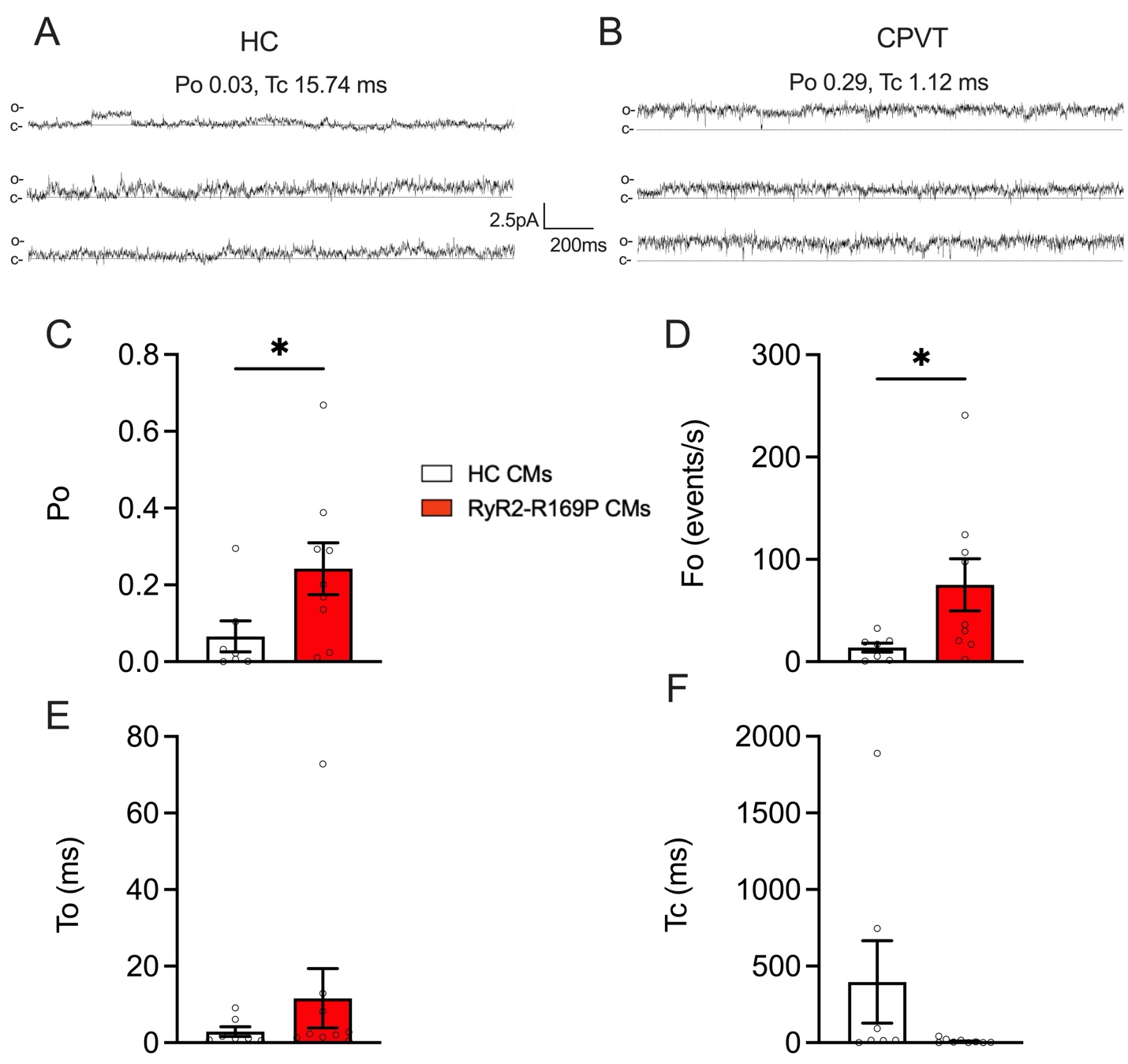

**Figure S5: (A and B)** Representative illustrations of PKA-phosphorylated RyR2 single channel recordings under 150 nmol/L free cis [Ca^2+^] in HC and CPVT hiPSC-CMs. The « c- » and « o- » represent the RyR2 channel closed and opened state, respectively. **(C)** Scatter plot with each data point superimposed showing the single channel open probability (Po) of RyR2 channels in HC and CPVT SR microsomes. **(D)** Frequency of openings (Fo in events/s) of RyR2 channels in HC and CPVT SR microsomes. **(E)** Mean open time (To in ms) of RyR2 channels in HC and CPVT SR microsomes. **(F)** Mean closed time (Tc in ms) of RyR2 channels in HC and CPVT SR microsomes. The number of experiments varies from 7 to 9 in HC and CPVT hiPSC-CM lysates. Data are presented as mean ± SEM.

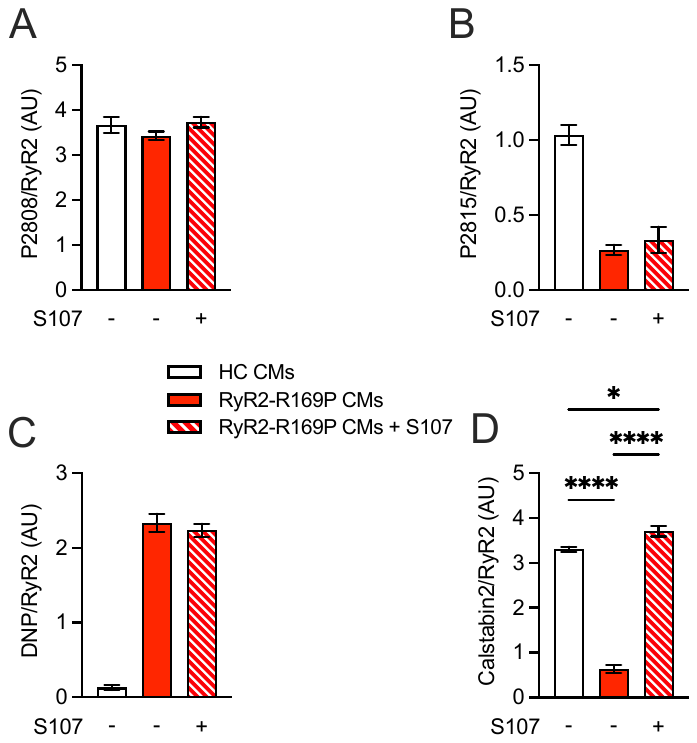

**Figure S6: (A)** Bar graphs showing the relative RyR2 PKA-phosphorylation level at Ser 2809 in HC and RyR2-R169P hiPSC-CMs +/- S107. **(B)** Relative RyR2 CaMKII-phosphorylation level at Ser 2815 in HC and RyR2-R169P hiPSC-CMs +/- S107. **(C)** Relative RyR2 oxidation level in HC and RyR2-R169P hiPSC-CMs +/- S107. **(D)** Relative calstabin2 amount bound to RyR2 in HC and RyR2-R169P hiPSC-CMs +/- S107. Data are presented as mean ± SEM.

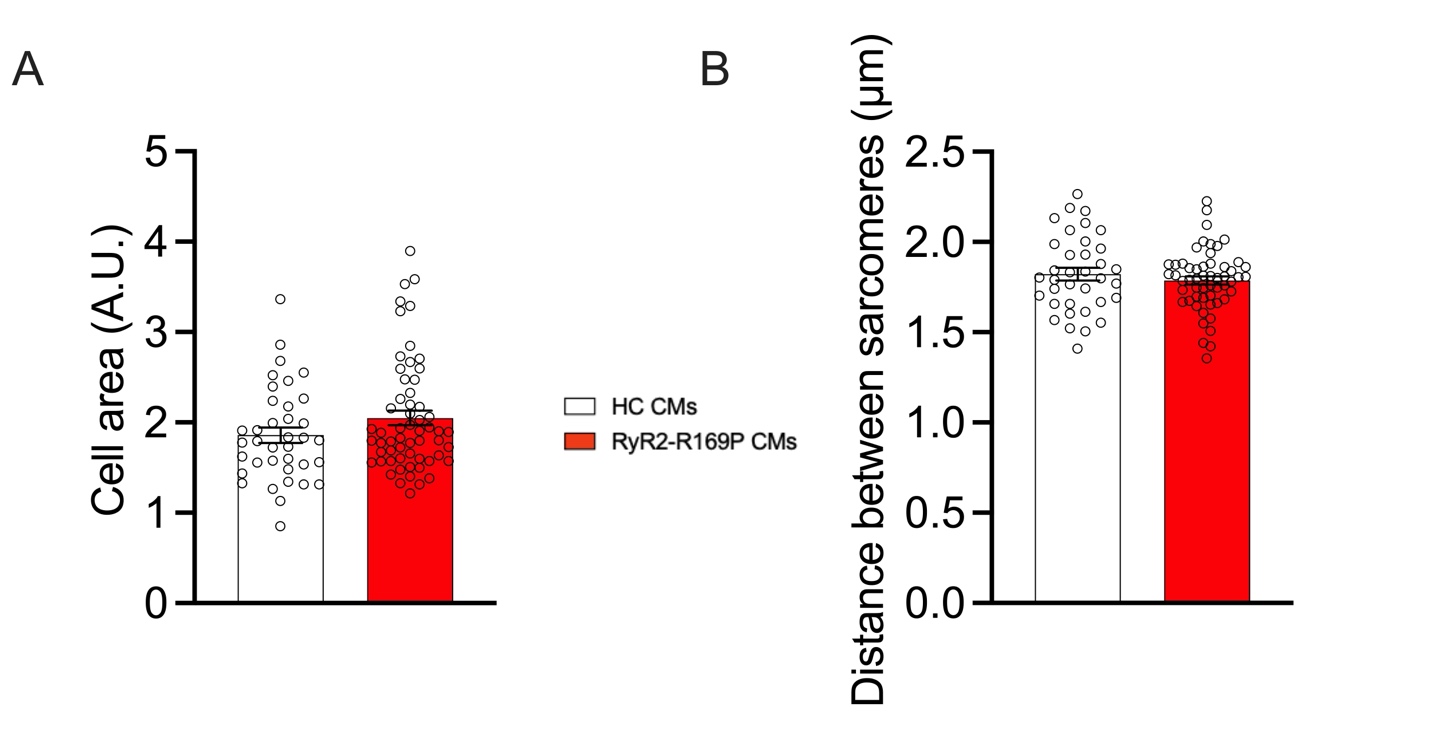

**Figure S7: (A)** Bar graphs showing the cell area in HC and RyR2-R169P hiPSC-CMs. **(B)** Bar graphs showing the distance between sarcomeres in HC and RyR2-R169P hiPSC-CMs. Data are presented as mean ± SEM.

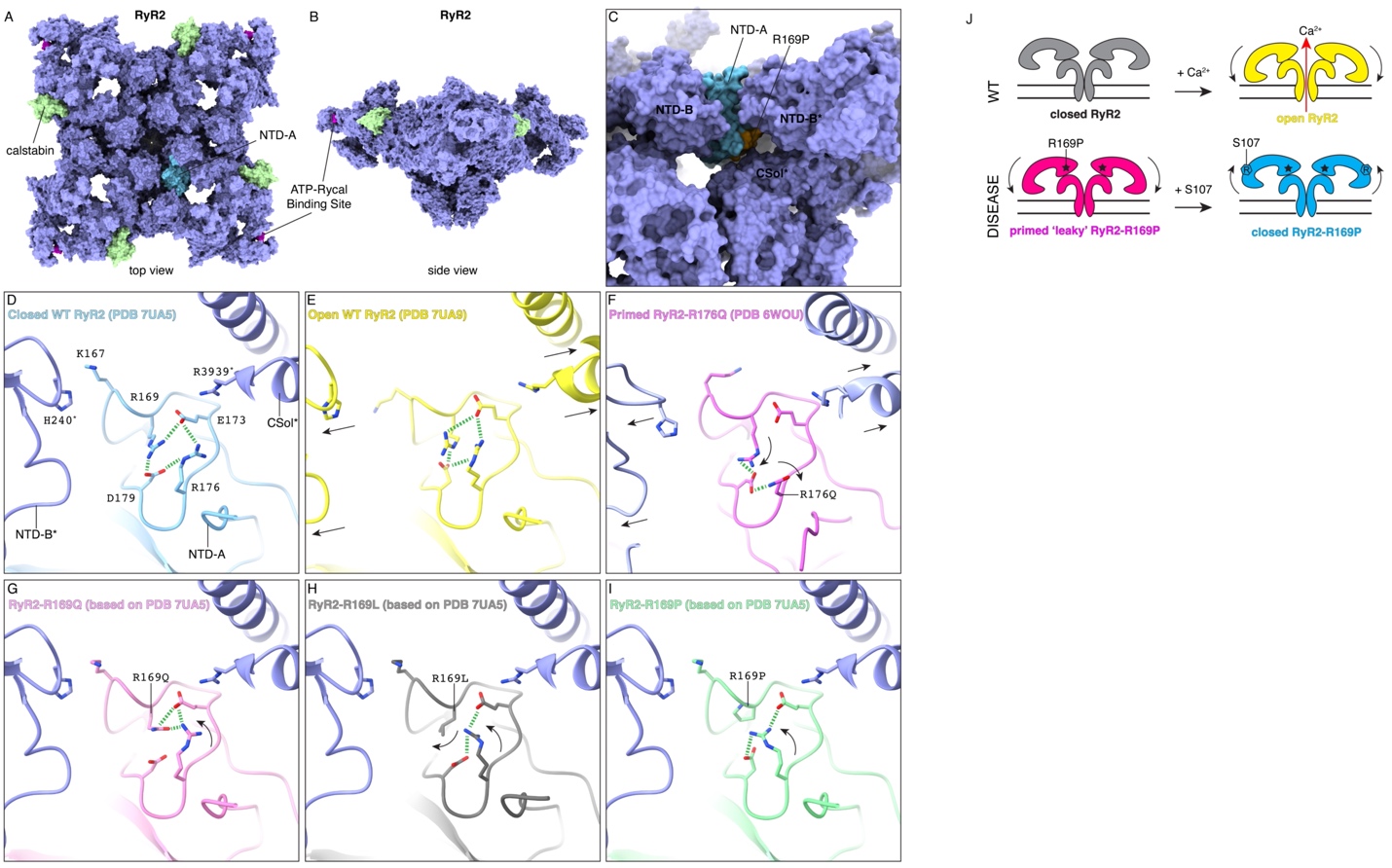

**Figure S8:** Localization and structural effects of RyR2-R169P mutation. (A,B) RyR2 atomic model (PDB 7UA5) showing the NTD-A domain (cyan) where R169P is located, as well as the ATP-Rycal binding site (magenta), and the stabilizing calstabin subunit (green). (C) Internal view of the RyR2 atomic model, showing the loop where R169P is located (orange), and how it interacts with the NTD-B* and CSol* domains of the neighbor protomer (*). (D) RyR2 atomic model (PDB 7UA5) showing the residues involved in the stabilization of the NTD-A loop (R169, E173, R176, and D179), and the residues involved in the interaction with the NTD-B* (K167 and H240*) and CSol* (G172 and R3939*) domains. (E) Same as (D) but with open RyR2 (PDB 7UA9). The interaction between the NTD-A and the NTD-B* and CSol* domains is disrupted leading to an outward shift of such domains (shown in arrows). (F) Same as (D) but with CPVT RyR2-R176Q (PDB 6WOU). Mutation R176Q disrupts the intra-NTD-A stabilizing interactions, leading to a flexibilization of the loop, loss of interaction with the NTD-B* and CSol* domains, outward shift of such domains (shown in arrows), and induction of the primed state. (G-I) Same as (D) but with modelled CPVT-related mutations RyR2-R169Q (G), RyR2-R169L (H), and RyR2-R169P (I). All three mutations would disrupt the intra-NTD-A stabilizing interactions, while R169Q would have similar effects to R176Q (both Arg to Gln mutations), R169L and R169P would have more pronounced effects because of hydrophobic repulsion (R169L and R169P) and backbone torsion (R169P). (J) WT RyR2 under resting conditions remains closed with the cytoplasmic shell in an upward-inward position, while in the presence of Ca^2+^, the cytoplasmic shell adopts a downward-outward position that together with Ca^2+^ binding leads to the pore opening. Disease-related RyR2-R169P, as other CPVT mutants, would be in a primed state, where the cytoplasmic shell is in a downward-outward position, lowering the energetic barrier to reach the open state, resulting in a ‘leaky’ channel. Binding of Rycal S107 induces an upward-inward shift of the cytoplasmic shell, preventing the ‘leak’ and restoring a normal function.

**Material and methods:**

***Ethics statement***

Written informed consent form was obtained from the parents of the minor patient with the RyR2-R169P mutation, who agreed to provide blood samples for hiPSC generation and basic research. This study was conducted in accordance with the Declaration of Helsinki and was approved by the Montpellier Hospital Review Board Committee (approval number 1003-HPS2). The male healthy control and RyR2-H29D and its isogenic control hiPSC lines used in this study were previously characterized ^1, 2^.

***3D in silico modeling***

3D *in silico* modeling was performed using the structures of human RyR2 in the closed (PDB: 7UA5) and open (PDB: 7UA9) states and of the mouse RyR2-R176Q (PDB: 6WOU). Visualization, *in silico* mutation, and hydrogen bond analysis was performed using the software ChimeraX ^3^.

***hiPSC generation and maintenance***

Following blood sample and PBMC isolation, we reprogrammed in bulk and fully characterized the hiPSC line from the CPVT patient harboring RyR2-R169P as previously described ^4^. The hiPSC were maintained on extracellular matrix Matrigel hES-Qualified and cultured into StemFlex culture media at 37°C in 5% O_2_ and 5% CO_2_.

***Cardiac differentiation protocol***

We carried out cardiac differentiation using the 2D cardiac sheet protocol, which modulates Wnt/β-catenin signaling pathway, using CHIR99021 and Wnt-C59 cytokines, as we have previously published ^1^*.* At day 20, the media RPMI/B27 was supplemented with 100nM of Triiodothyronine (T3) and 1µM of Dexamethasone to improve the ventricular-like hiPSC-CM phenotype ^5^. All experiments were done at day 30 of the cardiac differentiation.

***Cardiac sarcoplasmic reticulum (SR) vesicle preparation***

SR vesicles were isolated from hiPSC-CMs exposed to 1µM isoproterenol for 1 hour at 37°C to mimic the beta-adrenergic stimulation. The isolation of SR vesicles was performed as previously described to maintain the native and unpurified state of RyR2 ^2^.

***RyR2 single-channel recordings by the planar lipid bilayer technique***

SR vesicles with RyR2 were integrated into planar lipid bilayers by applying a lipid solution of asolectin (type IV, Sigma) at a concentration of 45mg/mL in decane at room temperature (RT), following established protocols ^2^. The properties of the channels, such as open probability (Po) and frequency of openings (Fo), were analyzed according to previously described methods ^6^.

***Neuronal differentiation protocol***

We carried out neuronal differentiation using the STEMdiff Midbrain Neuron Differentiation kits (#100-0038 and 100-0041, StemCell Technologies). The hiPSC were seeded on Matrigel hES-Qualified into Neural Induction Medium for 3 weeks, with a passage every 7 days. On day 21, the cells were dissociated and seeded in Neural Progenitors Medium expansion medium for 14 days, with a passage after 1 week. The neuronal progenitors thus obtained undergo another passage in Neural Differentiation Medium supplemented SHH cytokine (200ng/mL), for 1 week. Sonic Hedgehog (SHH) plays a major role in structuring the CNS ^7^. It acts directly on cells to specify the fate of neural cells, regulates the proliferation and survival of oligodendrocyte precursors and neural crest cells, and stimulates the proliferation, differentiation and growth of axons. The immature neurons were dissociated and seeded for 3 weeks in Neural Maturation Medium to obtain mature hiPSC-NRs.

***Immunocytochemistry (ICC)***

The hiPSC-CMs and -NRs were fixed in 4% paraformaldehyde (PFA) for 15 min at room temperature (RT) and rinsed 3 times with PBS. Cells were permeabilized in a solution of 0.1% triton in PBS for 10 min at RT. Then the cells were blocked with 0.1% bovine serum albumin (BSA) in PBS at RT for 30 min. The cardiac (a-actinin at 1:1000 and Troponin I at 1:1000) and the neuronal antibodies (ß3-tubulin at 1:1000) were diluted in a solution containing 0.1% BSA and PBS, were incubated for 2 hours at RT. The secondary antibodies, diluted in a solution containing 0.1% BSA and PBS and 40,6-diamidino-2-phenylindole (DAPI), were then incubated for 1 hour at RT. Cells were rinsed 3 times with PBS and stocked at 4°C. Immunocytochemistry was performed using the ZEISS LSM800 confocal microscope with the Z-stack option for 3D imaging. The resulting images were analyzed using ImageJ and previously published Morphoscript in Matlab ^8^.

***Quantitative measurement of the cytosolic calcium levels***

The hiPSC-CMs and -NRs were incubated with 1µM INDO-1 AM ratiometric dyes for 15 minutes at 37°C in Tyrode’s solution (1.8mM CaCl2, 10mM Glucose, 10mM HEPES, 4mM KCl, 1mM MgCl2, 135mM NaCl). Imaging was performed in Tyrode’s solution at RT using the IonOptix Myocam-S. Fluorescence was analyzed using IonWizard software.

***Fluorescent confocal microscopy and intracellular calcium handling***

The hiPSC-CMs and -NRs were incubated with 1µM Fluo-4 AM for 15 minutes at 37°C in culture media. The experiments were conducted in Tyrode’s solution (1.8mM CaCl2, 10mM Glucose, 10mM HEPES, 4mM KCl, 1mM MgCl2, 135mM NaCl) at 37°C. Ca^2+^ transients were captured using a ZEISS LSM780 confocal microscope. For the hiPSC-CMs, Ca^2+^ transients were recorded for 7 seconds in line scan mode and analyzed using PeakInspector as previously described ^9^. For hiPSC-NRs, frame mode was used during a 5-minute acquisition period and analyzed using a custom-developed MATLAB script ^10^.

***Neurotransmitter release quantification with mass spectrometry***

The cell secretion media were recovered, and cells were resuspended in 50µL of 0.1mM ascorbic acid before membrane disruption (4 × 10 s, 100 W; Fisher Scientific, Waltham, MA, USA; Model 505 Sonic Dismembrator). After centrifugation (20,000× g, 30 min, 4°C), supernatants were recovered and protein concentrations were determined (Protein Assay kit; Bio-Rad, Marnes-la-Coquette, France). We used the isotopic dilution method for absolute quantification ^11^. 20µL of the cell extracts or 50µL of the secretion medium were mixed with 10µL of internal standards containing 20pM of D4-dopamine, C6-noradrenaline, D6-adrenaline, D3-L-DOPA, 25pmole of D6-GABA and 90pmol of D5-glutamate in 0.1mM ascorbic acid. 40µL of borate buffer and 10µL AccQtag Ultra reagent (AccQ-Tag Ultra derivatization kit, Waters, Guyancourt, France) were added to the samples followed by a 10 min incubation at 55 °C under agitation. Samples were mixed with 500µL of ice-cold acetonitrile (ACN) and centrifuged (20,000×g, 30min, 4°C). Finally, the supernatants were dried under vacuum and resolubilized in 20µL of 0.1% formic acid (v/v).

Analyses were performed on a Dionex Ultimate 3000 HPLC system (Thermo Scientific, Waltham, MA, USA) coupled with an Endura triple quadrupole mass spectrometer (Thermo Electron, Waltham, MA, USA). The system was controlled by Xcalibur v. 4.0 software (Thermo Electron). Next, 5µL of samples were loaded into reverse phase Zorbax column C18-SB (#863600-902; 1mm × 150mm, 3.5µm, Agilent Technologies, Santa Clara, CA, USA, SB-C18). Elution of the compounds was performed at a flow rate of 90µL/min, at 40°C (see Table S1 for conditions). Buffer A corresponded to H2O 98.9%/ACN 1%/formic acid 0.1% (v/v/v) and buffer B was ACN 99.9%/formic acid 0.1% (v/v). Dopamine, adrenaline, noradrenaline, L-DOPA, GABA and glutamate were measured using the multiple reaction monitoring mode (MRM) according to the settings. The targeted compounds are detailed in Supplement Table S1. The selection of the monitored transitions and the optimization of the collision energy (CE) were manually determined. The identification of the compounds was based on precursor ions, daughter ions and retention times obtained for dopamine, adrenaline, noradrenaline, L-DOPA, GABA and glutamate and their corresponding internal standards. Amounts of neurotransmitters were quantified according to the isotopic dilution method ^11^.

***Immunoprecipitation and immunoblot analyses***

The hiPSC-CMs and -NRs were incubated with a lysis buffer containing 35mM NaF, 50mM Tris maleate (pH 6.8), 1mM Na_3_VO_4_, and protease inhibitors as previously published ^2^. RyR2 channels were immunoprecipitated by incubating 100mg of cell lysate with an anti-RyR antibody (homemade: rabbit 5029 y2) for 2 hours at 4°C in 0.5ml of RIPA buffer (10mM Tris-HCl, pH 7.4, 150mM NaCl, 5mM NaF, 1mM Na3VO4, 1% Triton-X100, and protease inhibitors). The immune complexes were then incubated with protein A sepharose beads (GE Healthcare, ref: 17528001) overnight at 4°C, followed by three washes with RIPA buffer. Proteins were separated using a 4-20% SDS-PAGE gradient gel, transferred onto nitrocellulose membranes, and incubated overnight at 4°C with primary antibodies: rabbit 5029 Y2 anti-RyR2 (1:5000), anti-phospho-RyR2-pSer2809 (homemade: polyclonal rabbit CRTRRI-(pS)-QTSQ, 1:1000), anti-RyR2-pSer2815 (homemade: polyclonal rabbit CSQTSQV-(pS)-VD), anti-Cys-NO (Sigma-Aldrich, 1:1000), anti-DNP (Millipore, 1:2000), tubulin (Abcam, ref: EPR13796), and mouse anti-FKBP12.6 (Santa Cruz, ref: 376135, 1:1000). Levels of RyR2-bound proteins were normalized to the total RyR2 immunoprecipitated (arbitrary units). All immunoblots were developed using the Odyssey system (LI-COR) with IR-labeled secondary antibodies (1:30,000 dilution) for 1 hour at RT.

***Teratoma* *formation***

We evaluated the pluripotency and differentiation potential of hiPSCs through teratoma formation. Healthy Control (HC) and CPVT hiPSCs were cultured and injected subcutaneously (3x10^6^ per line) into anesthetized NOD/SCID/J mice, as previously described ^4^. In some experiments, hiPSC were pretreated with S107 (5µM for 10 days in culture) before being injected into mice, which were then pretreated with S107 (20 mg/kg/day in the drinking water) until excision. After a latency period of 4–8 weeks, histological analysis was performed on the excised tumor nodules, which were fixed and embedded in paraffin. Sections of the paraffin blocks were placed on slides and stained with HES (Hemalun Eosin Saffron) to determine present structures derived from the three embryonic layers. Another serial section was stained with PAS (Periodic Acid-Schiff) to highlight glycosaminoglycans and complex carbohydrates, which appear fuchsia pink and are typical of Goblet cells, characteristic of the respiratory and digestive epithelium, both derived from the endoderm layer. The slides were scanned using the Hamamatsu MRI slide scanner (Hamamatsu Photonics, Massy, France) and analyzed with its NDPview software for image capture.

***Statistical analysis***

Normality was tested using the Shapiro-Wilk test. A t-Test was used to compare 2 independent groups with parametric distribution and a Mann-Whitney test was performed to compare 2 independent groups with non-parametric distribution. An ANOVA test was performed to compare 3 independent groups with parametric distribution and a Kruskal-Wallis was performed to compare 3 independent groups with non-parametric distribution. All data are expressed as mean±SEM. Data analysis and statistics were done with GraphPad Prism (version 9). *, p<0.05; **, p<0.01; ***, p< 0.001; ****, p< 0.0005.
